# Stain-robust classification of Alzheimer’s disease and chronic traumatic encephalopathy

**DOI:** 10.64898/2026.11.22.26368692

**Authors:** Lingyi Xu, Mark C. Nwakamma, Christopher Tilton, Bobak Abdolmohammadi, Ann C. McKee, Thor D. Stein, Jonathan D. Cherry, Jesse B. Mez, Vijaya B. Kolachalama

**Author notes:** Corresponding author: Vijaya B. Kolachalama, PhD.

## Abstract

Retrospective reassessment of archival brain banks provides opportunities to identify previously undocumented neuropathologies. However, limited tissue availability and preservation, together with differences in sampling and staining protocols across brain banks, can con-strain the histological information available for analysis. This study investigated whether computational models can integrate complementary information across histological stains and retain discriminative information under incomplete stain availability. A deep learning model incorporating shared-specific feature modeling, cross-stain knowledge distillation, and random modality masking was developed and evaluated on 1, 607 whole slide images from the dorsolateral frontal cortex of 602 donors across three cohorts. Using chronic traumatic encephalopathy (CTE) and Alzheimer’s disease (AD) and their comorbidity as a proof-of-principle application, our model achieved macro-averaged areas under the receiver operating characteristic curve of 0.899, 0.904, and 0.898 across three stain-defined groups, exceeding corresponding single-stain baselines. Discriminative performance was retained when each group’s defining stain was withheld. Model-predicted disease probabilities were associated with neuropathological staging measures for AD and CTE, and spatial attribution patterns on the remaining stains were largely preserved under defining-stain masking. These findings demonstrate the feasibility of leveraging complementary and partially overlapping histopathological information under incomplete stain availability, providing a foundation for future computational studies across brain regions and brain banks.

## Introduction

Brain banks are an important resource for investigating the pathological basis of neurodegenerative diseases. As diagnostic criteria evolve, retrospective assessment of archival brain tissue offers opportunities to identify pathological changes that may not have been evaluated during the original examination. Chronic traumatic encephalopathy (CTE), for which con-sensus neuropathological diagnostic criteria were established relatively recently, represents a particularly relevant example [1, 2]. Reassessment of existing brain banks for CTE is of particular interest given the prevalence of Alzheimer’s disease (AD) pathology in aging populations and the challenges associated with distinguishing these two diseases. AD and CTE can occur independently or coexist, and their shared involvement of hyperphosphory-lated tau pathology can complicate disease-specific interpretation, particularly in advanced or mixed-pathology cases [3, 4, 5]. Computational tools that facilitate the characterization of such complex neuropathological profiles could support retrospective brain bank research and assist pathologists in the evaluation of overlapping disease processes.

A practical challenge in retrospective neuropathological research is the availability of the histological information needed for disease characterization. Additional sectioning or staining of archival brain tissue may be constrained by tissue availability and preservation, particularly years after the original examination [6, 7, 8, 9]. Furthermore, differences in tis-sue sampling and staining protocols across brain banks can result in systematic differences in the brain regions and histological preparations available for analysis [10]. These challenges are relevant to the study of neurodegenerative diseases, for which pathological assessment relies on multiple complementary histological measurements. In AD and CTE, hyperphos-phorylated tau pathology, commonly visualized using antibodies such as AT8, is central to CTE diagnosis and contributes to AD staging [11, 1]; amyloid-*β* (A*β*) immunohistochemistry reveals amyloid plaque pathology [12]; and Luxol fast blue combined with hematoxylin and eosin (LHE) provides structural and myelin context. While these stains highlight distinct pathological and morphological features, they may also contain partially overlapping information relevant to disease characterization. Computational approaches capable of leveraging this complementarity without requiring uniform stain availability could facilitate the use of heterogeneous brain bank resources.

Whole slide imaging (WSI) has enabled high-resolution digitization of histological preparations and the development of computational approaches ranging from multiple instance learning to graph- and transformer-based aggregation [13, 14, 15, 16, 17, 18, 19]. How-ever, most computational neuropathology pipelines remain stain-specific and focused on individual diseases or lesion types [20], while multimodal approaches often assume complete modality availability during training and inference [21, 22, 23, 24]. Existing strategies for missing-modality learning, including imputation, generative synthesis and modality-specific cascades, have been investigated primarily in radiological imaging [25, 26], with limited ex-ploration in multi-stain WSI analysis. An important question is whether computational models can effectively integrate complementary information across histological stains and retain discriminative information when individual stains are unavailable.

This study addresses this question using AD and CTE classification as a proof-of-principle application within a single brain region. Although comprehensive neuropathological assess-ment of AD and CTE involves the evaluation of multiple brain regions [27, 1], restricting the anatomical scope provides a setting in which to investigate cross-stain information integra-tion. A multimodal deep learning model was developed to classify cases into four mutually exclusive categories: AD, CTE, comorbid AD and CTE, and neither. Building on the shared-specific feature-modeling paradigm for missing-modality learning [28], the model combines stain-specific transformer encoders with a shared encoder aligned through cross-modal knowl-edge distillation and employs random modality masking during training to accommodate absent stains at inference. The framework was developed and evaluated on 1, 607 WSIs from 602 donors spanning three neuropathology cohorts: the Understanding Neurologic Injury and Traumatic Encephalopathy (UNITE) brain bank, the Framingham Heart Study (FHS), and the Boston University Alzheimer’s Disease Research Center (ADRC), each contributing AT8-, A*β*-, and LHE-stained sections of the dorsolateral frontal cortex, a region heavily affected in CTE and routinely sampled for neuropathological assessment of AD [3, 27]. To evaluate the benefits of multimodal integration and its robustness to missing stains, three overlapping study groups were defined, each using a different stain (AT8, A*β*, or LHE) as its defining stain. Within each group, a unimodal model using only the defining stain was compared with a multimodal model incorporating all available stains from the same cases. The multimodal model was further evaluated with the defining stain withheld to assess the extent to which predictive information could be retained through the remaining stains. This design enables matched comparisons across different single-stain baselines without privileging any particular stain as a universally required input. Associations between model-predicted disease proba-bilities and neuropathological staging measures for AD and CTE, as well as the stability of spatial attribution under defining-stain masking, were additionally evaluated to characterize model behavior beyond classification performance. Together, these investigations establish a basis for future computational neuropathology studies integrating histological information across multiple brain regions and brain banks with heterogeneous sampling and staining protocols.

## Materials and methods

### Study population

Whole slide images (WSIs) obtained from the dorsolateral frontal cortex region were analyzed from three brain banks focusing on AD and CTE pathology (Figure 1A): the Understanding Neurologic Injury and Traumatic Encephalopathy (UNITE), the Framingham Heart Study (FHS), and Boston University Alzheimer’s Disease Research Center (ADRC). The dorsolateral frontal cortex was selected on the basis of its involvement as one of the earliest and most severely affected cortical regions in CTE and its inclusion among the standard sampling regions for neuropathological assessment of AD [3, 27]. The three datasets comprised WSIs derived from formalin-fixed paraffin-embedded (FFPE) brain tissue samples, with staining protocols including AT8 for hyperphosphorylated tau, A*β* for amyloid-beta (A*β*) plaques, and Luxol-fast blue combined with hematoxylin and eosin (LHE) for general tissue morphology. The Boston University ADRC longitudinally tracks older research participants across the cognitive spectrum, from normal cognition to dementia, and includes diagnostic slides digitized at 20× magnification [29, 30]. The FHS employs identical staining and digitization protocols as the ADRC and is a multi-generational, community-based longitudinal cohort affiliated with the same brain bank [31]. The UNITE Brain Bank serves as a repository for individuals with histories of repetitive head impacts, such as from contact sports or military service [32]. The UNITE samples included in this study were selected with an emphasis on high-stage CTE to facilitate comparative histopathological analysis of AD and CTE. Intake and processing protocols for these cohorts were standardized across all brain banks, with neuropathology assessments performed by board-certified neuropathologists using the NIA-Reagan criteria for AD [33], and the second NINDS/NIBIB consensus criteria for CTE [1]. Both 20× and 40× stained WSIs were included. Cases were included if they had at least two different stains out of AT8, A*β*, and LHE, and no neurodegenerative pathologies other than AD and/or CTE. Other neurodegenerative pathologies were excluded to minimize potential confounding from overlapping histopathological features. This yielded a total of 602 cases (UNITE: *n* = 418, FHS: *n* = 83, ADRC: *n* = 101) and 1, 607 WSIs (AT8: *n* = 512, A*β*: *n* = 519, LHE: *n* = 576). Stain availability varied across cases, resulting in heterogeneous combinations of histological stains within the study population. Summary statistics and clinical features are reported in Table 1.

**Figure 1.**
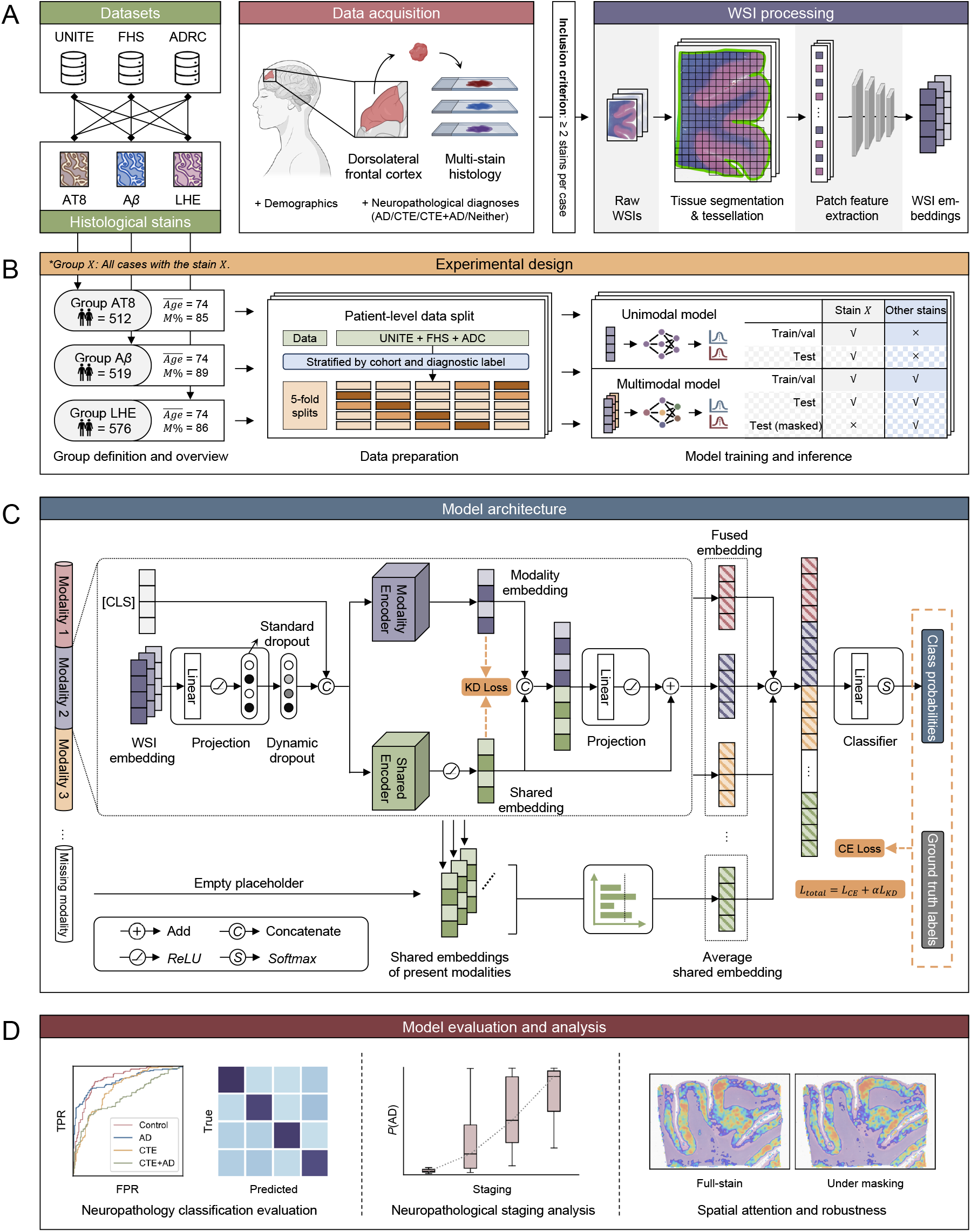
Data, experimental design, model architecture, and evaluation. **A.** Whole-slide images (WSIs) of three histological stains (AT8, A*β*, and LHE) from the dor-solateral frontal cortex are obtained from three independent cohorts (UNITE, FHS, and ADRC). Cases with at least two stains available are included. WSIs undergo tissue segmen-tation and tessellation, followed by patch-level feature extraction to generate WSI embed-dings. **B.** Three overlapping stain-defined groups are constructed to evaluate the benefits of multimodal integration relative to different single-stain baselines. Each group includes all cases with its defining stain (AT8, A*β*, or LHE). Within each group, the unimodal model uses only the defining stain, whereas the multimodal model uses all available stains from the same cases, enabling matched comparisons without differences in case composition. Cases from all three cohorts are pooled and divided into five folds, jointly stratified by cohort and diag-nostic label. In each cross-validation iteration, three folds (60%) are used for training, one (20%) for validation, and one (20%) for testing, with validation and test assignments rotated so that each case serves in each role exactly once. Stochastic modality masking is applied during multimodal training. Each multimodal model is additionally evaluated with its defin-ing stain withheld at inference to assess robustness under systematic stain absence. **C.** The multimodal framework comprises three modality-specific transformer encoders and a shared transformer encoder for learning stain-dependent and shared representations, respectively. The resulting representations are integrated within each stain and concatenated across stains to form a case-level embedding. Missing-stain embeddings are substituted with the average shared embedding from available stains. A linear classification head followed by softmax pro-duces probabilities for four mutually exclusive diagnostic categories (Control, AD, CTE, and CTE+AD). Training combines classification supervision with shared-specific representation alignment. **D.** Models are evaluated using held-out test predictions from five-fold cross-validation, with classification metrics computed both per fold and from pooled out-of-fold predictions. Evaluation includes four-class classification performance, associations between predicted disease probabilities and neuropathological staging measures for AD and CTE, characterization of stain-specific spatial attention patterns, and spatial attribution stability under defining-stain masking.

**Table 1:**
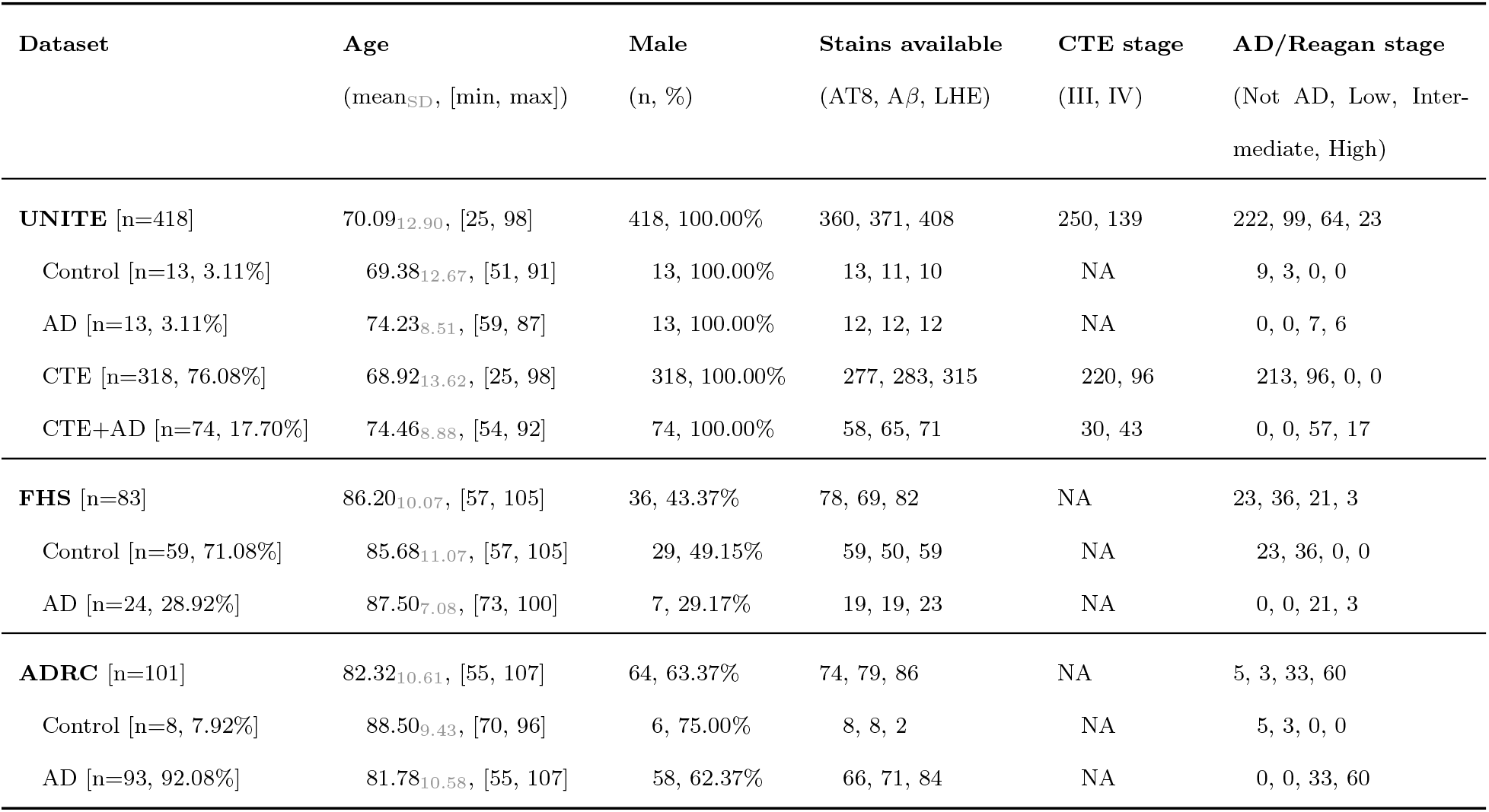
Study population. Summary of demographic and clinical attributes for the three datasets used in the study. Each dataset section reports the mean age (with the SD as a gray subscript and the [min, max] range; 3 UNITE/CTE cases with missing age excluded from the age statistic only), number of male subjects (with percentage), the availability of WSI stains (AT8, A*β*, and LHE), the CTE stage breakdown (Stage III and Stage IV), and the AD/Reagan stage breakdown (Not AD, Low Probability, Intermediate Probability, High Probability). Staging counts may not sum to the total number of cases due to missing staging information.

| **Dataset** | **Age** | **Male** | **Stains available** | **CTE stage** | **AD/Reagan stage** |
|---|---|---|---|---|---|
|  | (mean<sub>SD</sub>, [min, max]) | (n, %) | (AT8, Aβ, LHE) | (III, IV) | (Not AD, Low, Inter-mediate, High) |
| **UNITE** [n=418] | 70.09<sub>12.90</sub>, [25, 98] | 418, 100.00% | 360, 371, 408 | 250, 139 | 222, 99, 64, 23 |
| Control [n=13, 3.11%] | 69.38<sub>12.67</sub>, [51, 91] | 13, 100.00% | 13, 11, 10 | NA | 9, 3, 0, 0 |
| AD [n=13, 3.11%] | 74.23<sub>8.51</sub>, [59, 87] | 13, 100.00% | 12, 12, 12 | NA | 0, 0, 7, 6 |
| CTE [n=318, 76.08%] | 68.92<sub>13.62</sub>, [25, 98] | 318, 100.00% | 277, 283, 315 | 220, 96 | 213, 96, 0, 0 |
| CTE+AD [n=74, 17.70%] | 74.46<sub>8.88</sub>, [54, 92] | 74, 100.00% | 58, 65, 71 | 30, 43 | 0, 0, 57, 17 |
| **FHS** [n=83] | 86.20<sub>10.07</sub>, [57, 105] | 36, 43.37% | 78, 69, 82 | NA | 23, 36, 21, 3 |
| Control [n=59, 71.08%] | 85.68<sub>11.07</sub>, [57, 105] | 29, 49.15% | 59, 50, 59 | NA | 23, 36, 0, 0 |
| AD [n=24, 28.92%] | 87.50<sub>7.08</sub>, [73, 100] | 7, 29.17% | 19, 19, 23 | NA | 0, 0, 21, 3 |
| **ADRC** [n=101] | 82.32<sub>10.61</sub>, [55, 107] | 64, 63.37% | 74, 79, 86 | NA | 5, 3, 33, 60 |
| Control [n=8, 7.92%] | 88.50<sub>9.43</sub>, [70, 96] | 6, 75.00% | 8, 8, 2 | NA | 5, 3, 0, 0 |
| AD [n=93, 92.08%] | 81.78<sub>10.58</sub>, [55, 107] | 58, 62.37% | 66, 71, 84 | NA | 0, 0, 33, 60 |

### Diagnostic criteria

Neuropathological diagnoses of AD and CTE followed consensus criteria. AD pathology was assessed using the National Institute on Aging-Reagan criteria [33], which combine Braak staging of neurofibrillary tangle topography [11] with CERAD scoring of neuritic plaque density [34] to estimate the likelihood that AD pathology accounts for dementia. In this study, AD neuropathology was categorized into four Reagan categories (none, low, inter-mediate, and high). CTE was diagnosed according to the second NINDS/NIBIB consensus criteria [1], which require a characteristic perivascular accumulation of hyperphosphorylated tau in neurons, with or without glial involvement, at the depths of cortical sulci. CTE neuropathology was assessed using the McKee staging scheme, which defines four stages (I-IV) based on the extent and anatomical distribution of tau pathology [3]. Stages I-II and III-IV broadly correspond to low and high CTE staging, respectively, as described in the second NINDS/NIBIB consensus recommendations [1]. Cases meeting criteria for both AD and CTE were adjudicated as comorbid (CTE+AD). All diagnoses were rendered by board-certified neuropathologists blinded to clinical data and brain bank membership, following the standardized intake and staging protocols shared across UNITE, FHS and ADRC.

### WSI processing

An existing image segmentation pipeline was used on LHE WSIs to isolate tissue from background and artifacts [35]. For AT8- and A*β*-stained WSIs, dedicated tissue segmen-tation models were trained per stain using available gray-matter region annotations. Gray matter annotations were matched to corresponding WSIs, yielding 252 AT8 and 218 A*β* annotation-WSI pairs, split case-wise into training and hold-out test sets at a 4:1 ratio (seed 42), producing 203*/*49 cases for AT8 and 175*/*43 for A*β*. WSIs were downsampled to 10× magnification and annotation polygons were rasterized as binary tissue masks at the same resolution. Tiles of 512×512 pixels were extracted with stride 512 and a minimum tissue fraction of 0.15. A U-Net [36] with a ResNet-18 encoder pretrained on ImageNet was trained for binary tissue-versus-background segmentation. Training used a combined binary cross-entropy and Dice loss, AdamW optimizer (learning rate 0.0001, one-cycle schedule), and early stopping with patience of 10 on the Dice score on the validation set. Segmentation performance is reported in Supplemental Figure S1. For study population cases with existing gray matter annotations (324 AT8 and 279 A*β* slides), annotations were used directly as tissue masks. For the remaining cases without annotations (188 AT8 and 240 A*β* slides), trained UNet models were applied to generate tissue masks.

Following tissue segmentation, tiles were extracted without overlap at 512×512 pixels for 20× WSIs and 1024×1024 pixels for 40× WSIs. Each tile was resized to 448×448 pixels and passed through a patch encoder pretrained on over 350k diverse H&E and IHC slides [37], producing a 1536-dimensional embedding ***f*** ∈ R^1×1536^ used as input to the downstream model.

### Modeling framework

The multimodal model was built upon the shared-specific representation learning framework introduced in ShaSpec [28]. It consists of a multimodal backbone that integrates information across stains and a linear classification head that predicts probabilities of each of four mutually exclusive diagnostic categories: Control, AD, CTE, or comorbid CTE+AD (Figure 1C). The backbone comprises three modality-specific transformer encoders, one for each stain (AT8, A*β*, and LHE), and a shared transformer encoder with weights tied across stains. All encoders follow the TransMIL architecture [18], consisting of two Nystromformer attention layers with pyramidal positional encoding between them. This architecture was selected for its effectiveness in WSI multiple-instance learning and its computationally efficient attention mechanism for processing large numbers of image patches.

For each available stain, WSI patch embeddings were projected into the transformer input space through a linear layer and processed with a custom input-dependent patch dropout module. During training, patches were ranked by the sigmoid-transformed mean of their embedding features and assigned nonlinearly increasing dropout probabilities, up to a maxi-mum of *p*. Selected patches were stochastically zeroed to introduce nonuniform regularization and reduce reliance on high-scoring patch representations. Dropout was disabled during inference. The modality-specific encoders learned stain-dependent representations, while the shared encoder learned representations across stains through shared parameters. For each stain, the modality-specific and shared embeddings were concatenated and passed through a linear projection followed by a ReLU activation. A residual connection from the shared embedding was then added to produce an integrated stain representation. The integrated embeddings from all three stain positions were concatenated into a case-level representation and passed through a linear classification head, followed by softmax to obtain four-class probabilities. When a stain was unavailable, its integrated embedding was replaced by the average of the shared embeddings from the available stains, preserving a fixed-dimensional input to the classifier regardless of stain availability.

Training was guided by a composite objective combining four-class classification super-vision with cross-modality representation alignment. Given the imbalance among the four mutually exclusive diagnostic categories (Tables 1, Supplemental Table S1), classification supervision used a class-weighted cross-entropy loss. For predicted class logits ***x*** ∈ R^4^ and ground-truth label *y* ∈ {0, 1, 2, 3}, the classification loss was defined as:

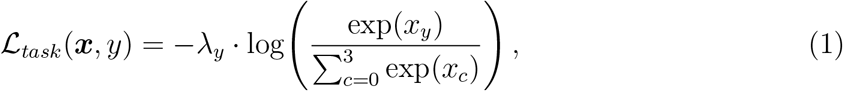

where class weights *λ_c_* = *N/N_c_* were calculated from the training-set class frequencies, with *N* denoting the total number of training cases and *N_c_* the number belonging to class *c*. In addition, a class-balanced random sampler was used during training to reduce imbalance in the frequency of diagnostic categories encountered during optimization.

To align shared and modality-specific representations, a knowledge distillation [38] (KD) objective was applied at the representation level using the L1 distance. For each case, the KD loss was calculated as the sum of the alignment losses across all available stains:

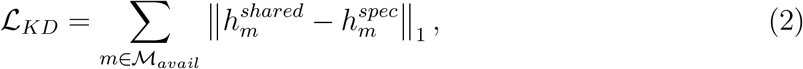

where M*_avail_* denotes the set of available stains, and *h^shared^* and *h^spec^* are the outputs of the shared and corresponding modality-specific encoders for stain *m*, respectively. Missing stains were excluded from the alignment loss. This objective encourages the shared representations to retain information captured by the stain-specific encoders. The total training loss combined classification and representation alignment:

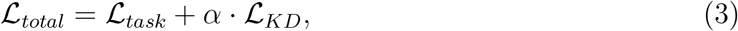

where *α* controls the relative contribution of representation alignment to the training objec-tive.

For comparison, a unimodal model was trained separately for each stain-defined group us-ing only its defining stain. Each unimodal model employed the same TransMIL-based trans-former architecture as the modality-specific encoder in the multimodal backbone, followed by the same linear four-class classification head. Optimization was achieved by minimizing the class-weighted cross-entropy classification objective L*_task_*.

### Experimental design

To systematically evaluate the benefits of multimodal integration relative to different single-stain baselines, three overlapping groups were defined within the study population based on stain availability (Figure 1B): Group AT8 (*n* = 512) included all cases with AT8, Group A*β* (*n* = 519) included all cases with A*β*, and Group LHE (*n* = 576) included all cases with LHE. Models were trained and evaluated separately for each stain-defined group. Within each group, the unimodal model used only the defining stain, whereas the multimodal model incorporated all available stains for the same cases. This design enabled paired comparisons between unimodal and multimodal models while controlling for differences in case composition. Repeating these comparisons across three defining stains allowed the bene-fits of multimodal integration to be evaluated without assuming any particular stain as a universally required input.

To improve robustness to incomplete stain availability, stochastic modality masking was applied during multimodal training. Each available stain was independently masked with a probability of 0.3, with masks resampled whenever all available stains were selected for re-moval so that at least one stain remained per case. Masked stains were represented by empty embeddings. In addition to full-input evaluation, each multimodal model was evaluated un-der a defining-stain-masked condition, in which the group’s defining stain was withheld for every test case. This condition assessed the extent to which predictive performance could be retained through the remaining stains under systematic absence of the defining stain.

For each stain-defined group, cases from all three cohorts (UNITE, FHS, and ADRC) were pooled and evaluated using stratified five-fold cross-validation. Splits were constructed at the case level, jointly stratified by cohort and diagnostic label to preserve cohort-specific diagnostic composition across folds. In each iteration, three folds (60%) were used for train-ing, one fold (20%) for validation, and one fold (20%) for testing. Validation and test assignments were rotated across the five iterations so that each case served as a validation case exactly once and as a held-out test case exactly once. Unimodal and multimodal models were trained independently for each iteration, with early stopping and checkpoint selection based on validation-set macro-averaged four-class AUROC. The same data partitions were used for unimodal and multimodal models within each group to enable paired comparisons. Model performance was evaluated both within individual held-out test folds and using pooled out-of-fold predictions across all five iterations, as detailed in Performance metrics.

Models were trained using the AdamW optimizer with a learning rate of 0.0002 and weight decay of 0.0001. Standard dropout used a dropping probability of 0.25, and input-dependent patch dropout used a maximum dropping probability of 0.3. The transformer hidden dimension was set to 128, and the knowledge distillation loss weight was set to *α* = 0.2 for multimodal training. Training was performed for up to 200 epochs, with early stopping patience of 20 epochs. Batch size of 4 was used for all three stain groups based on hyperparameter tuning, with gradient accumulation applied to accommodate memory constraints. Class-balanced oversampling was applied during training to mitigate diagnostic label imbalance. A fixed random seed of 2025 was used for reproducibility.

### Performance metrics

Models were evaluated as four-class classifiers over the mutually exclusive diagnostic categories (Control, AD, CTE, and CTE+AD). Classification performance was assessed using the area under the receiver operating characteristic curve (AUROC), average precision (AP), accuracy, precision, sensitivity, and specificity. AUROC and AP were calculated separately for each class as a one-vs-rest problem against the remaining three classes, then macro-averaged across all four classes to obtain the overall (All) scores. Accuracy, precision, sensitivity, and specificity were computed from the argmax predictions of the four-class softmax out-puts. Precision, sensitivity, and specificity were macro-averaged across the four one-vs-rest confusion matrices derived from these predictions.

Performance was evaluated at the individual-fold and pooled out-of-fold levels. For each cross-validation iteration, all classification metrics were calculated independently on the corresponding held-out test set to characterize fold-level performance. In addition, classification metrics were calculated on a pooled out-of-fold evaluation population constructed by concatenating held-out predictions from all five iterations. The pooled evaluations were used for the primary model comparisons and statistical analyses, ensuring that results were not dependent on any single test fold.

Model comparisons were performed separately within each stain-defined group using differences in pooled macro-averaged four-class AUROC (ΔAUROC). Three pairwise comparisons were evaluated: multimodal versus unimodal, defining-stain-masked multimodal versus unimodal, and defining-stain-masked multimodal versus multimodal. Standard errors and 95% confidence intervals were estimated using 10,000 class-stratified bootstrap resamples of the pooled test population. The 95% confidence intervals were obtained from the 2.5th and 97.5th percentiles of the bootstrap distributions. Two-sided *p*-values were calculated using Wald tests based on the ratio of the observed ΔAUROC to its bootstrap standard error and the standard normal approximation. Combined estimates were obtained by averaging the three stain groups’ ΔAUROC values and their corresponding bootstrap resample distributions with equal weight per group. Bootstrap resampling was coordinated at the donor level across the three overlapping stain groups to preserve dependencies arising from shared cases. Holm-Bonferroni correction [39] was applied across the three combined model com-parisons, while group-level *p*-values were reported without adjustment. All statistical tests were two-sided with a significance threshold of 0.05.

For the neuropathological staging analyses, the predicted probability of AD pathology, *P* (AD), was defined as the sum of the softmax probabilities assigned to the AD and CTE+AD classes, while the predicted probability of CTE pathology, *P* (CTE), was defined as the sum of the probabilities assigned to the CTE and CTE+AD classes. These probabilities cap-tured disease-specific predictions regardless of comorbidity. Associations between predicted disease probabilities and neuropathological staging categories were assessed using Spearman rank-order correlations. AD probabilities were evaluated across four Reagan categories (no AD, low, intermediate and high), while CTE probabilities were evaluated between cases without CTE and those with high-stage CTE (stages III-IV). Differences in predicted prob-abilities between staging categories were assessed using two-sided Mann-Whitney *U* tests, with Holm-Bonferroni correction across the six pairwise comparisons within each AD analy-sis. All staging analyses were conducted separately for each stain-defined group and model condition using pooled out-of-fold test predictions. Cases with missing staging information were excluded from the corresponding staging analyses.

### Spatial interpretation and masking-invariance analysis

Two types of attention-derived maps were extracted from the trained multimodal models to characterize spatial attention and class-specific attribution. First, modality-specific encoder attention maps were obtained by extracting the CLS-row attention vectors from each trans-former layer, averaging across attention heads within each layer, and then averaging across the two layers. The resulting map assigned one attention weight to each patch, reflecting its relative contribution to attention-based representation aggregation. Second, shared-encoder class attribution maps were computed using a gradient-weighted attention method [40]. For each case, the attribution target was the logit corresponding to its diagnostic label (Con-trol, AD, CTE, or CTE+AD). At each transformer layer *l*, the gradient of the target-class logit *ℓ* with respect to the CLS-row attention vector *A*^(*l*)^ was obtained by backpropagation.

Per-layer attribution was calculated as

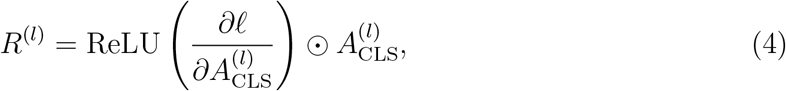

where ⊙ denotes element-wise multiplication. Attribution values were averaged across attention heads and then across the two transformer layers to obtain a per-patch attribution map. This approach retains positively contributing attention gradients and weights them by attention magnitude using additive aggregation across layers.

For representative case visualization, cases were selected from the held-out test samples of the median-performing cross-validation fold, identified by the mean four-class macro-AUROC across the three stain groups. Attention and attribution maps were then extracted using the corresponding fold-specific models. Per-patch values were mapped to downsampled WSI thumbnails using patch coordinates retained during preprocessing. Following the CLAM visualization convention [41], values were converted to percentile ranks, with the lower half suppressed and the upper half linearly rescaled to [0, 1]. Rasterized maps were smoothed using a Gaussian kernel with *σ* = max(8 px, 0.6 × *w_p_*), where *w_p_* denotes patch width in thumbnail pixels. Smoothing was restricted to regions covered by actual patches using a hard coverage mask. The resulting maps were overlaid on WSI thumbnails with an opacity of 0.40 using a truncated jet colormap excluding the upper 20% of the standard color range to reduce visual overlap with the brown DAB chromogen in AT8 and A*β* slides. Modality-specific attention maps for AT8 and A*β* were restricted to the gray-matter annotation polygons used during patch extraction; LHE maps were not spatially restricted to gray matter.

To quantify spatial attribution stability under missing-stain conditions, Spearman rank-order correlations (*ρ*) were calculated between full-input and defining-stain-masked shared-encoder attribution vectors before rasterization or visualization normalization. The analysis included cases with all three stains available, using pooled out-of-fold evaluations across all five cross-validation folds. For each case within a stain-defined group, correlations were computed separately for the two non-defining stains, comparing corresponding patch-level attribution values under full and masked inference. Both conditions used the case’s diagnostic label as the attribution target, yielding one correlation per case–non-defining stain combination. Correlations were summarized by stain group, non-defining stain, and diagnostic label, with an additional overall summary pooling all observations across the three groups.

Statistical evidence of high attribution consistency was assessed using one-sided, one-sample Wilcoxon signed-rank tests [42] of whether the correlations exceeded a reference threshold of *ρ* = 0.8. Tests were conducted separately for each group–non-defining stain–ground-truth label combination and for the pooled overall distribution. Holm-Bonferroni correction [39] was applied across all tests to control the family-wise error rate, with adjusted *p <* 0.05 considered statistically significant.

### Computational hardware and software

The model implementation and all training and inference procedures were developed in Python 3.11.10, using PyTorch 2.7.0 and PyTorch Lightning 2.5.1. Other supporting Python libraries include pandas 2.0.3, NumPy 1.23.5, TorchMetrics 1.7.1, scikit-learn 1.2.1, SciPy 1.14.1, and Matplotlib 3.7.1. Training and inference were conducted on a single NVIDIA L40S GPU on a shared computing cluster.

### Data availability

Data from UNITE, FHS and ADRC cohorts may be requested through the respective data cores, subject to institutional approval.

### Code availability

Python scripts and trained model weights will be made available on GitHub upon acceptance of the manuscript.

## Results

Four-class classification of Control, AD, CTE, and CTE+AD was evaluated within three overlapping stain-defined groups (AT8, A*β*, and LHE; Figure 1). Within each group, the unimodal model used only the defining stain, whereas the multimodal model incorporated all available stains from the same cases. The multimodal model was additionally evaluated with the defining stain withheld at inference. Cases from the three cohorts were pooled within each group and evaluated using five-fold cross-validation, with case-level splits stratified jointly by cohort and diagnostic label. All reported results were obtained from held-out test data, including pooled out-of-fold predictions for aggregate analyses. Cohort-level demographics and stain availability are summarized in Table 1. Demographics, stain availability, and cohort composition for each stain-defined group are presented in Supplemental Table S1.

### Multimodal classification performance

Each multimodal model was evaluated as a single four-class classifier over mutually exclu-sive diagnostic categories using pooled out-of-fold predictions spanning the study population of each stain-defined group (512 cases for AT8, 519 for A*β*, 576 for LHE). Overall macro-averaged classification AUROC reached 0.899 for AT8, 0.904 for A*β*, and 0.898 for LHE (Table 2). Per-class ROC and PR curves are shown in Figure 2A-F, and row-normalized con-fusion matrices are shown in Figure 2G-I. Across all three stain groups, the model achieved consistently high per-class AUROCs, ranging from 0.852 to 0.946, indicating broadly effec-tive discrimination across the four diagnostic categories. Per-fold performance metrics were reported in Supplemental Table S2.

**Figure 2.**
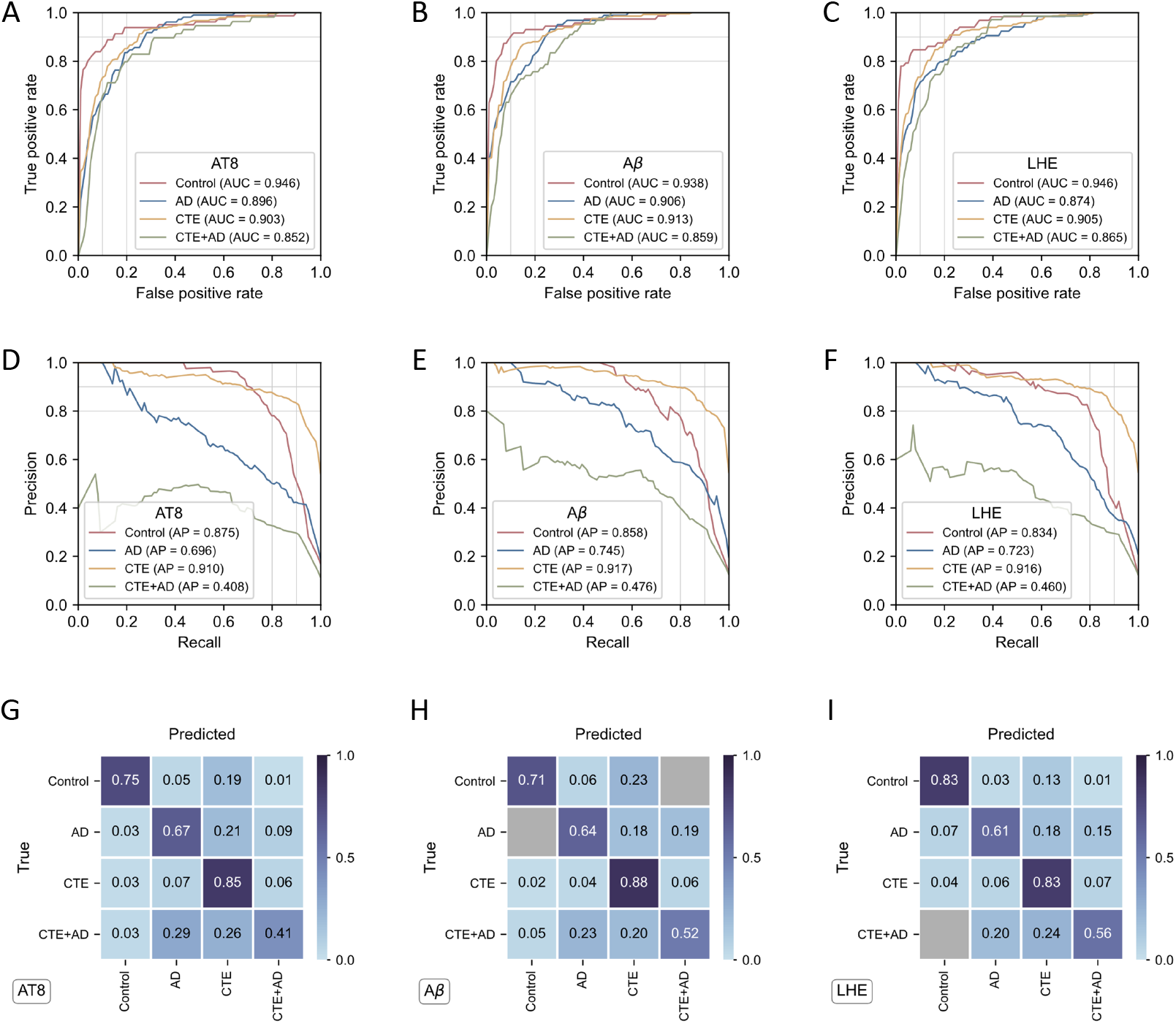
Performance on four-class classification. Results are shown for the full-input multimodal models using pooled out-of-fold test predictions across all five cross-validation folds for each stain group. **A-C.** Receiver operating characteristic (ROC) curves for the AT8 (**A**), A*β* (**B**), and LHE (**C**) groups. Each diagnostic class (Control, AD, CTE, and CTE+AD) is evaluated as a one-vs-rest classification problem against the remaining three classes. The area under the ROC curve (AUROC) is reported in the legend. **D-F.** Precision-recall (PR) curves for the same stain groups and diagnostic classes, with average precision (AP) reported in the legend. **G-I.** Row-normalized confusion matrices for the AT8 (**G**), A*β* (**H**), and LHE (**I**) groups, based on argmax predictions from the four-class softmax outputs. Each cell represents the proportion of cases belonging to the true class (row) predicted as the corresponding class (column). Gray cells indicate zero entries.

**Table 2:**
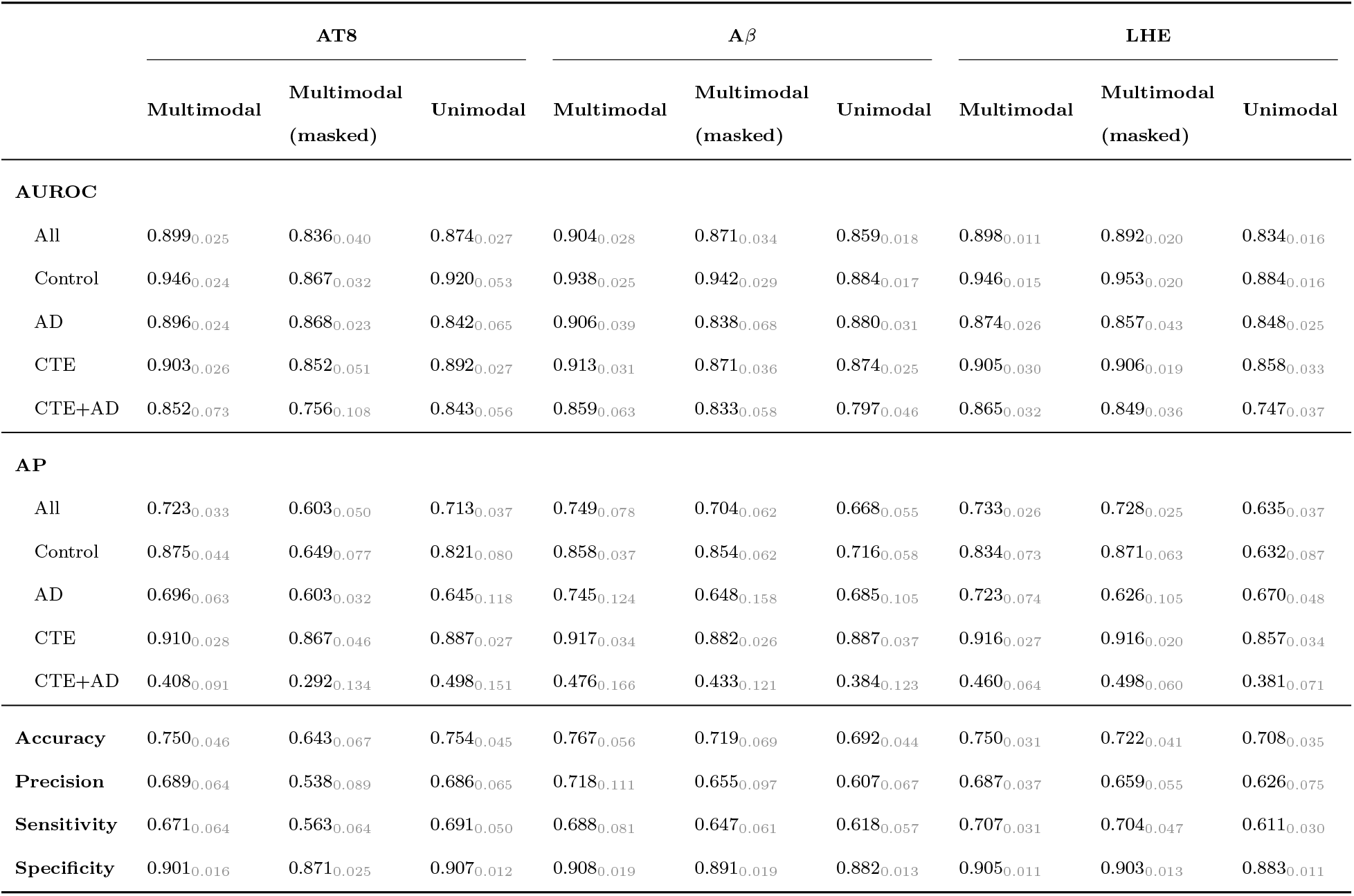
Model performance on four-class (Control / AD / CTE / CTE+AD) classification. For each defining stain (AT8 / A*β* / LHE), unimodal and multimodal classifiers are evaluated using stratified five-fold cross-validation, with each case contributing to the held-out test set in exactly one fold. Unimodal models use only the defining stain, whereas multimodal models use all available stains and are additionally evaluated with the defining stain masked at inference. Each cell reports the metric calculated from pooled out-of-fold predictions across all five folds, with the SD (gray subscript) calculated from fold-specific held-out test metrics. AUROC and AP are reported for each class using a one-vs-rest formulation and as macro-averages across the four classes (All). Accuracy, precision, sensitivity, and specificity were calculated from arg-max predictions over the four diagnostic classes; precision, sensitivity, and specificity were macro-averaged across classes.

|  | **AT8** |  |  | **Aβ** |  |  | **LHE** |  |  |
|---|---|---|---|---|---|---|---|---|---|
|  | **Multimodal** | **Multimodal<br>(masked)** | **Unimodal** | **Multimodal** | **Multimodal<br>(masked)** | **Unimodal** | **Multimodal** | **Multimodal<br>(masked)** | **Unimodal** |
| **AUROC** |  |  |  |  |  |  |  |  |  |
| All | 0.899<sub>0.025</sub> | 0.836<sub>0.040</sub> | 0.874<sub>0.027</sub> | 0.904<sub>0.028</sub> | 0.871<sub>0.034</sub> | 0.859<sub>0.018</sub> | 0.898<sub>0.011</sub> | 0.892<sub>0.020</sub> | 0.834<sub>0.016</sub> |
| Control | 0.946<sub>0.024</sub> | 0.867<sub>0.032</sub> | 0.920<sub>0.053</sub> | 0.938<sub>0.025</sub> | 0.942<sub>0.029</sub> | 0.884<sub>0.017</sub> | 0.946<sub>0.015</sub> | 0.953<sub>0.020</sub> | 0.884<sub>0.016</sub> |
| AD | 0.896<sub>0.024</sub> | 0.868<sub>0.023</sub> | 0.842<sub>0.065</sub> | 0.906<sub>0.039</sub> | 0.838<sub>0.068</sub> | 0.880<sub>0.031</sub> | 0.874<sub>0.026</sub> | 0.857<sub>0.043</sub> | 0.848<sub>0.025</sub> |
| CTE | 0.903<sub>0.026</sub> | 0.852<sub>0.051</sub> | 0.892<sub>0.027</sub> | 0.913<sub>0.031</sub> | 0.871<sub>0.036</sub> | 0.874<sub>0.025</sub> | 0.905<sub>0.030</sub> | 0.906<sub>0.019</sub> | 0.858<sub>0.033</sub> |
| CTE+AD | 0.852<sub>0.073</sub> | 0.756<sub>0.108</sub> | 0.843<sub>0.056</sub> | 0.859<sub>0.063</sub> | 0.833<sub>0.058</sub> | 0.797<sub>0.046</sub> | 0.865<sub>0.032</sub> | 0.849<sub>0.036</sub> | 0.747<sub>0.037</sub> |
| **AP** |  |  |  |  |  |  |  |  |  |
| All | 0.723<sub>0.033</sub> | 0.603<sub>0.050</sub> | 0.713<sub>0.037</sub> | 0.749<sub>0.078</sub> | 0.704<sub>0.062</sub> | 0.668<sub>0.055</sub> | 0.733<sub>0.026</sub> | 0.728<sub>0.025</sub> | 0.635<sub>0.037</sub> |
| Control | 0.875<sub>0.044</sub> | 0.649<sub>0.077</sub> | 0.821<sub>0.080</sub> | 0.858<sub>0.037</sub> | 0.854<sub>0.062</sub> | 0.716<sub>0.058</sub> | 0.834<sub>0.073</sub> | 0.871<sub>0.063</sub> | 0.632<sub>0.087</sub> |
| AD | 0.696<sub>0.063</sub> | 0.603<sub>0.032</sub> | 0.645<sub>0.118</sub> | 0.745<sub>0.124</sub> | 0.648<sub>0.158</sub> | 0.685<sub>0.105</sub> | 0.723<sub>0.074</sub> | 0.626<sub>0.105</sub> | 0.670<sub>0.048</sub> |
| CTE | 0.910<sub>0.028</sub> | 0.867<sub>0.046</sub> | 0.887<sub>0.027</sub> | 0.917<sub>0.034</sub> | 0.882<sub>0.026</sub> | 0.887<sub>0.037</sub> | 0.916<sub>0.027</sub> | 0.916<sub>0.020</sub> | 0.857<sub>0.034</sub> |
| CTE+AD | 0.408<sub>0.091</sub> | 0.292<sub>0.134</sub> | 0.498<sub>0.151</sub> | 0.476<sub>0.166</sub> | 0.433<sub>0.121</sub> | 0.384<sub>0.123</sub> | 0.460<sub>0.064</sub> | 0.498<sub>0.060</sub> | 0.381<sub>0.071</sub> |
| **Accuracy** | 0.750<sub>0.046</sub> | 0.643<sub>0.067</sub> | 0.754<sub>0.045</sub> | 0.767<sub>0.056</sub> | 0.719<sub>0.069</sub> | 0.692<sub>0.044</sub> | 0.750<sub>0.031</sub> | 0.722<sub>0.041</sub> | 0.708<sub>0.035</sub> |
| **Precision** | 0.689<sub>0.064</sub> | 0.538<sub>0.089</sub> | 0.686<sub>0.065</sub> | 0.718<sub>0.111</sub> | 0.655<sub>0.097</sub> | 0.607<sub>0.067</sub> | 0.687<sub>0.037</sub> | 0.659<sub>0.055</sub> | 0.626<sub>0.075</sub> |
| **Sensitivity** | 0.671<sub>0.064</sub> | 0.563<sub>0.064</sub> | 0.691<sub>0.050</sub> | 0.688<sub>0.081</sub> | 0.647<sub>0.061</sub> | 0.618<sub>0.057</sub> | 0.707<sub>0.031</sub> | 0.704<sub>0.047</sub> | 0.611<sub>0.030</sub> |
| **Specificity** | 0.901<sub>0.016</sub> | 0.871<sub>0.025</sub> | 0.907<sub>0.012</sub> | 0.908<sub>0.019</sub> | 0.891<sub>0.019</sub> | 0.882<sub>0.013</sub> | 0.905<sub>0.011</sub> | 0.903<sub>0.013</sub> | 0.883<sub>0.011</sub> |

Consistent class-specific differences emerged across all three stain groups. Control and CTE generally achieved higher AUROCs and APs, whereas CTE+AD showed the lowest performance, particularly in AP. The confusion matrices revealed a similar pattern: all four diagnostic classes had their highest prediction proportions along the diagonal, although CTE+AD showed lower correct classification proportions and greater confusion with AD and CTE across all three stain groups. Separating comorbid CTE+AD from either disease alone remained particularly challenging.

### Multimodal integration improves predictive performance

The multimodal model consistently outperformed its unimodal counterpart in macro-averaged four-class AUROC across all three stain groups (Table 2). AUROC increased from 0.874 to 0.899 for AT8, from 0.859 to 0.904 for A*β*, and from 0.834 to 0.898 for LHE. Across the three groups, multimodal integration significantly improved four-class discrimination (ΔAUROC = 0.044, adjusted *p <* 0.0001; Supplemental Table S3). Consistent improvements were also observed in overall AP and precision across all three groups, and in accuracy, sensitivity, and specificity for A*β* and LHE (Table 2). At the individual class level, multimodal integration improved AUROC in all 12 stain-group/class comparisons and AP in 11 of 12 comparisons, with the only decrease in AP observed for CTE+AD in the AT8 group.

The magnitude of improvement varied across stain groups, with larger gains observed for stains with weaker unimodal discrimination. The smallest gain occurred in the AT8 group (ΔAUROC = 0.025, *p* = 0.029), followed by A*β* (ΔAUROC = 0.045, *p <* 0.0001), while LHE showed the largest improvement (ΔAUROC = 0.063, *p <* 0.0001; Supplemental Table S3). These findings demonstrate that multimodal integration provides benefits beyond individual stains, with the greatest observed gain in the LHE group, where the unimodal model had the lowest baseline performance.

### Multimodal models retain performance under missing-stain condi-tions

To evaluate robustness under systematic stain absence, each multimodal model was evaluated with its group’s defining stain withheld for every test case. This removed 35.6% (AT8), 35.9% (A*β*), and 37.0% (LHE) of the total WSIs available to each group’s multimodal model. Despite this reduction in available inputs, the masked multimodal models maintained macro-averaged four-class AUROCs of 0.836 for AT8, 0.871 for A*β*, and 0.892 for LHE (Table 2).

Compared with the corresponding unimodal baselines, masked multimodal performance was lower for AT8 (ΔAUROC = −0.038, *p* = 0.019), not different for A*β* (ΔAUROC = 0.013, *p* = 0.397), and higher for LHE (ΔAUROC = 0.057, *p <* 0.001; Supplemental Table S3).

The combined comparison across the three groups showed no differences between masked multimodal and unimodal performance (Supplemental Table S3). These results indicate that the ability to maintain discrimination using the remaining stains varies across stain groups, with masked multimodal performance approaching or exceeding the corresponding unimodal baseline for A*β* and LHE, though not for AT8.

Relative to the full multimodal models, withholding the defining stain reduced macro-averaged AUROC by varying magnitudes across the three groups. The largest reduction was observed when AT8 was withheld (ΔAUROC = −0.063, *p <* 0.0001), followed by A*β* (ΔAUROC = −0.033, *p <* 0.0001), whereas withholding LHE resulted in a smaller, non-significant reduction (ΔAUROC = −0.006, *p* = 0.328). Across the three groups, masking significantly reduced discrimination overall (ΔAUROC = −0.034, adjusted *p <* 0.0001; Supplemental Table S3). Together, these findings demonstrate that the impact of missing stains on predictive performance is not uniform, with the greatest observed dependence on AT8, intermediate dependence on A*β*, and minimal performance loss when LHE is withheld.

### Predicted disease probabilities are associated with neuropatholog-ical staging

To examine whether model predictions reflected neuropathological characteristics beyond categorical diagnostic labels, predicted disease probabilities were evaluated against established staging measures. For AD, predicted probabilities were compared across Reagan categories, derived from Braak neurofibrillary tangle stage and CERAD neuritic plaque score [11, 34, 33]. Median predicted AD probabilities increased monotonically across Reagan strata in all three stain groups (Figure 3A-C). Spearman rank-order correlations were 0.686 for AT8, 0.714 for A*β*, and 0.702 for LHE (all *p <* 0.0001), consistently exceeding the corresponding unimodal correlations of 0.607, 0.665, and 0.584, respectively (Supplemental Figure S2A-C). The largest increase in correlation was observed in the LHE group. Pairwise Mann-Whitney comparisons further showed significant differences in predicted probabilities across all six pairs of Reagan strata for both multimodal and unimodal models in every group (Holm-Bonferroni-adjusted *p <* 0.05).

**Figure 3.**
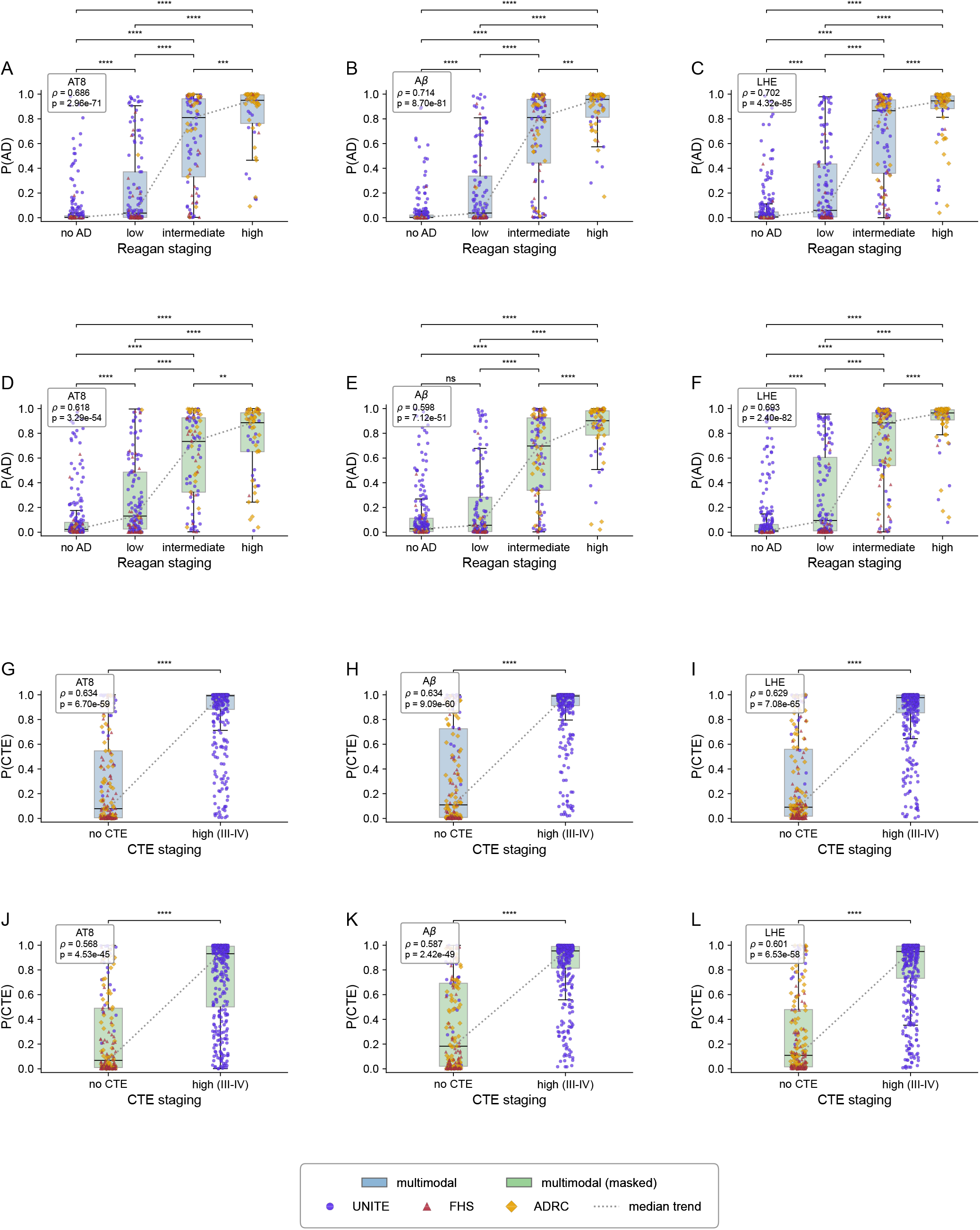
Model-predicted AD and CTE probabilities across neuropathologi-cal staging categories. Per-case predicted probabilities of AD and CTE are shown using pooled out-of-fold test predictions from all five cross-validation folds. **A-C.** Predicted AD probabilities across Reagan categories (no AD, low, intermediate, and high) from the full multimodal models for the AT8 (**A**), A*β* (**B**), and LHE (**C**) groups. **D-F.** Corresponding AD probabilities from the multimodal models evaluated with each group’s defining stain masked at inference for AT8 (**D**), A*β* (**E**), and LHE (**F**). **G-I.** Predicted CTE probabili-ties for cases without CTE and those with CTE (stages III-IV) from the full multimodal models for AT8 (**G**), A*β* (**H**), and LHE (**I**). **J-L.** Corresponding CTE probabilities under defining-stain masking for AT8 (**J**), A*β* (**K**), and LHE (**L**). Within each panel, individual cases are represented by scatter points (circles: UNITE; triangles: FHS; diamonds: ADRC). Boxplots summarize the distribution within each staging category (center line: median; box: interquartile range; whiskers extend to the most extreme observations within 1.5× the in-terquartile range), and dotted gray lines connect the category medians. Box colors indicate the input condition (blue: full multimodal; green: defining-stain-masked multimodal). Dif-ferences between staging categories are assessed using two-sided Mann-Whitney *U* tests, with Holm-Bonferroni correction across the six pairwise Reagan-category comparisons within each AD panel (A-F). CTE panels (G-L) compare the two available CTE categories. Significance is indicated by ns for adjusted *p* ≥ 0.05, ** for *p <* 0.01, *** for *p <* 0.001, and **** for *p <* 0.0001 (adjusted *p*-values for A-F). Spearman rank-order correlation coefficients (*ρ*) and their corresponding *p*-values are displayed in the upper-left corner of each panel.

To determine whether the association between predicted AD probabilities and neu-ropathological severity was maintained under missing-stain conditions, the analysis was repeated with each group’s defining stain masked at inference (Figure 3D-F). Spearman correlations declined from 0.686 to 0.618 for AT8, from 0.714 to 0.598 for A*β*, and from 0.702 to 0.693 for LHE. Pairwise Reagan stratum comparisons remained significant after masking, except for the no AD versus low comparison in the A*β* group. The smallest reduction was observed in LHE, consistent with its minimal loss in four-class discrimination under masking. Together, these results demonstrate that the association between predicted AD probabilities and neuropathological severity largely persists under missing-stain conditions, although its strength varies with the withheld stain.

CTE neuropathological staging is commonly categorized as low stage (I-II) and high stage (III-IV) [1]. The UNITE samples included in this study were selected with an emphasis on high-stage CTE (stages III-IV) to facilitate comparative histopathological analysis of AD and CTE. Predicted CTE probabilities were significantly higher in the high-stage group across all three stain groups (Figure 3G-I), with Spearman correlations of 0.634 for AT8, 0.634 for A*β*, and 0.629 for LHE (all *p <* 0.0001). These correlations exceeded the corresponding unimodal values of 0.614, 0.575, and 0.568 (Supplemental Figure S2D-F). Under defining-stain masking, the associations remained significant, with correlations of 0.568, 0.587, and 0.601 for AT8, A*β*, and LHE, respectively (Figure 3J-L). Thus, predicted CTE probabilities distinguished high-stage CTE from the absence of CTE across all model conditions.

### Spatial attention across stains and spatial attribution under defining-stain masking

Attention-derived maps were examined to characterize the spatial regions emphasized by the multimodal models and assess how these patterns changed when the defining stain was withheld. One representative case was selected from each stain group (AT8, A*β*, and LHE), corresponding to Case 1 (CTE+AD), Case 2 (AD), and Case 3 (CTE), respectively. Two complementary visualization approaches were used: modality-specific encoder attention maps and shared-encoder class attribution maps. Modality-specific attention maps are shown in Figure 4A. AT8 and A*β* attention maps highlighted regions consistent with the focal distribution of pathological features targeted by these stains. In contrast, LHE attention was more broadly distributed along the cortical gray matter, frequently forming continuous bands. These patterns suggest that the modality-specific encoders emphasize different histological features across stains.

**Figure 4.**
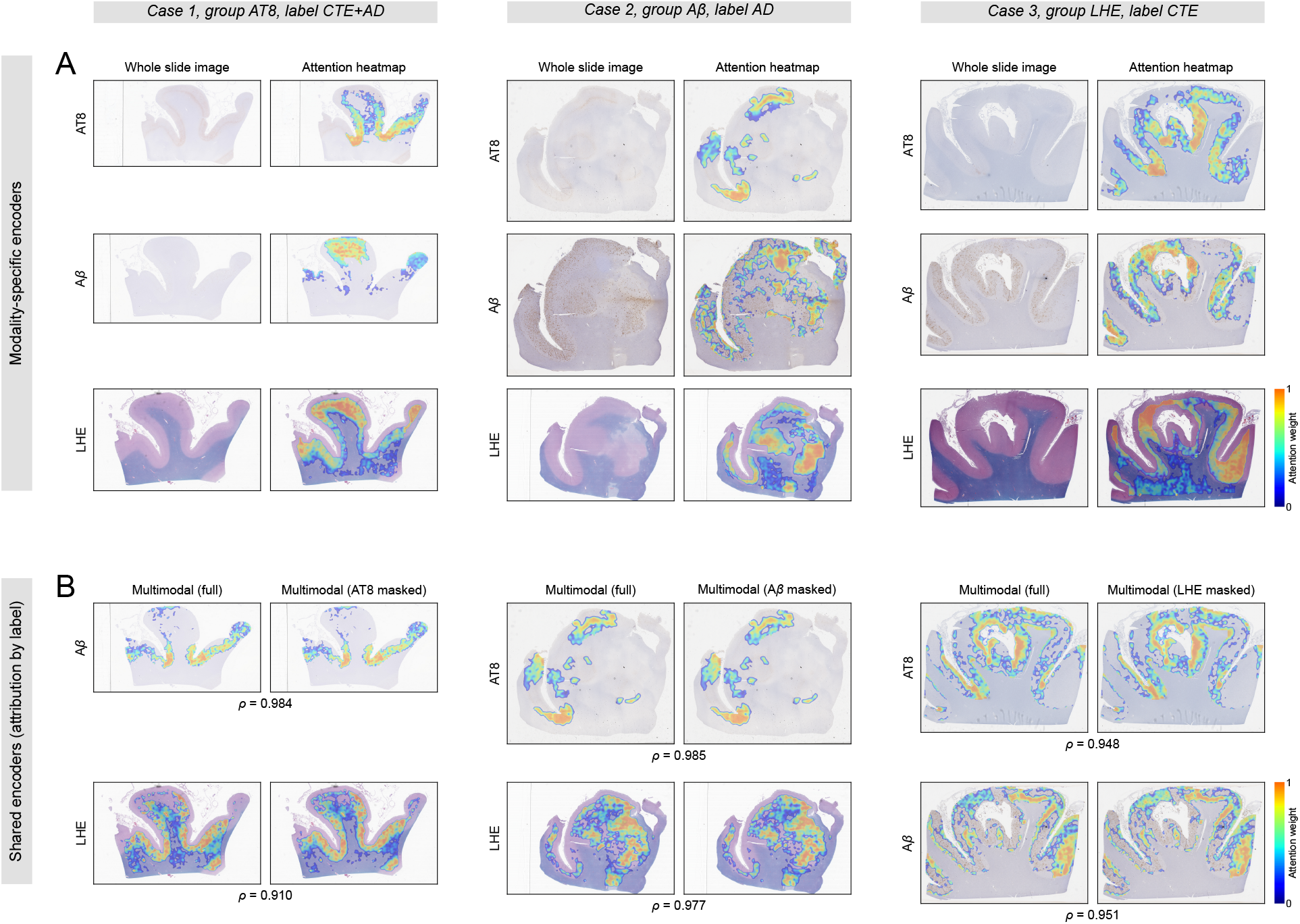
Spatial interpretation and missing-stain robustness. Three representative cases with all three stains available are shown, one from each stain-defined group: Case 1 (CTE+AD) from the AT8 group, Case 2 (AD) from the A*β* group, and Case 3 (CTE) from the LHE group. Representative cases are selected from the held-out test set of the median-performing cross-validation fold, determined by the mean four-class macro-AUROC across the three stain groups. **A.** Modality-specific encoder attention maps. For each case, whole-slide image thumbnails and their corresponding attention maps are displayed for AT8, A*β*, and LHE in separate rows. **B.** Shared-encoder attribution maps under defining-stain mask-ing. For each case, attribution maps are shown for the two non-defining stains, computed using gradient-weighted attention with the case’s true diagnostic label as the attribution tar-get. Within each case–stain pair, the left panel shows the multimodal model evaluated with all available stains, while the right panel shows the same model evaluated with the group’s defining stain masked. Spearman rank correlation (*ρ*) between the full and masked attri-bution vectors is reported below each masked panel. Population-level correlations across all four ground-truth diagnostic categories, restricted to cases with all three stains available, are reported in Supplemental Table S5. Colorbars indicate normalized attention or attribution weights on a [0, 1] scale.

To assess the stability of spatial attribution under missing-stain conditions, shared-encoder attribution maps, computed using each case’s true diagnostic label, were compared between full multimodal and defining-stain-masked inference for the two remaining stains in each group (Figure 4B). Per-case Spearman rank-order correlations between the corresponding full and masked attribution vectors were shown for each comparison, ranging from 0.910 to 0.985 across the three representative cases, indicating substantial preservation of spatial attribution patterns despite the removal of the defining stain. This finding was further sup-ported by a population-level analysis of all cases with three stains available, using pooled out-of-fold predictions across the five cross-validation folds. Across 2,418 case–stain–label combinations, the full-versus-masked attribution correlations averaged 0.887 ± 0.120 (median 0.927; Supplemental Table S5). One-sided Wilcoxon signed-rank tests against a correlation threshold of *ρ* = 0.8 showed evidence of correlations exceeding this threshold in every group–non-defining stain–label combination and in the pooled analysis (Holm-Bonferroni-adjusted *p <* 0.05). Together, these results demonstrate that the multimodal models largely preserve their spatial attribution patterns on the remaining stains when the defining stain is withheld, complementing the prediction-level robustness observed in the preceding analyses.

## Discussion

This study demonstrates the feasibility of integrating complementary histopathological information under incomplete stain availability for joint characterization of AD and CTE within a single brain region. Across three overlapping stain-defined populations, multimodal integration consistently improved four-class discrimination compared with matched unimodal baselines, while defining-stain masking demonstrated that predictive information could be retained through the remaining stains. Model-predicted disease probabilities were associated with neuropathological staging measures for AD and CTE, and spatial attribution patterns on the remaining stains were largely preserved under masking. Together, these findings support the integration of complementary neuropathological information without requiring complete stain panels.

The performance gains from multimodal integration and the effects of defining-stain masking revealed a consistent but asymmetric pattern across stain groups. AT8 showed the strongest unimodal discrimination and the smallest gain from multimodal integration, yet withholding AT8 caused the largest reduction in multimodal performance. In contrast, LHE showed the weakest unimodal discrimination and the largest benefit from additional stains, while withholding LHE had little effect on full multimodal performance. A*β* occupied an intermediate position in both comparisons. These observations suggest that the contribution of a stain within a multimodal framework depends not only on its individual discriminative capacity but also on the extent to which relevant information can be complemented by or recovered from other stains. The sensitivity to AT8 removal is biologically plausible given the central role of hyperphosphorylated tau in CTE diagnosis [1] and AD neurofibrillary pathology [11]. AT8 directly visualizes tau pathology that is not equivalently measured by A*β* or LHE, whereas LHE primarily provides structural and morphological information that may overlap with tissue features represented in the other stains. The smaller performance loss upon A*β* removal suggests that some information relevant to AD classification remains accessible through tau pathology and tissue morphology, although these stains do not directly measure amyloid plaques. These findings demonstrate that multimodal integration can leverage complementary information across stains to improve disease discrimination while exploiting partially overlapping information to mitigate the impact of missing stains.

Despite strong overall discrimination, comorbid CTE+AD remained the most challenging diagnostic category, with predictions frequently assigned to either CTE or AD alone. Unlike single-pathology classification, identifying comorbidity requires sufficient evidence for both pathological processes, which may differ in burden, spatial distribution, and detectability within the sampled tissue. CTE is characterized by focal perivascular hyperphosphorylated tau pathology at cortical sulcal depths [1], whereas AD neuropathological assessment incorporates neurofibrillary tangles and amyloid plaque pathology [11, 34, 33]. The difficulty of recognizing CTE+AD may therefore reflect incomplete or imbalanced evidence for the two disease components. Because neuropathological assessment of AD and CTE considers the distribution of pathology across multiple brain regions, the restriction to dorsolateral frontal cortex in this study may have limited the evidence available to distinguish these disease components in comorbid cases [27, 1]. These findings illustrate that multimodal integration within a single brain region can address only part of the complexity of neuropathological classification and underscore the importance of extending this approach to integrate information across anatomically distinct regions.

Methodologically, this work addresses a setting that differs from conventional single-stain computational pathology and multimodal fusion of histology with non-imaging data. Existing WSI classification approaches commonly aggregate patch-level information from individual stains through attention-based multiple-instance learning, graph-based modeling, or transformer architectures [13, 43, 41, 18, 19]. Other multimodal pathology frameworks combine histology with molecular or clinical measurements [22, 44, 45, 46]. Incomplete-modality learning has also been investigated in medical imaging through approaches such as HeMIS [47], SMIL [48], and ShaSpec [28], while emerging multi-stain pathology frameworks have begun addressing variable stain availability. Building on ShaSpec’s shared-specific

representation learning and missing-modality feature substitution, this study adapts these mechanisms to gigapixel multi-stain WSIs using TransMIL-based encoders for slide-level representation learning. The three stain-defined comparisons provide a systematic evaluation of multimodal integration relative to different single-stain baselines while controlling for differences in case composition within each comparison. Furthermore, withholding the defining stain for every test case evaluates model robustness under systematic stain absence, a condition relevant to differences in staining protocols across brain banks. This experimental framework provides a basis for evaluating multimodal integration under varying patterns of histopathological information availability.

Although the models were trained using categorical diagnostic labels, their predicted probabilities showed associations with neuropathological staging measures. For AD, predicted probabilities showed strong ordinal associations with Reagan neuropathological categories. Across all stain groups, full multimodal predictions were more strongly correlated with Reagan categories than their unimodal counterparts, with the largest improvement observed for LHE. Importantly, the predicted probability of AD incorporates AD-only and CTE+AD class probabilities, allowing the analysis to reflect AD-related pathology regard-less of CTE comorbidity. Because Reagan categories are derived from Braak neurofibrillary tangle stage and CERAD neuritic plaque score [11, 34, 33], the observed association is consistent with the models capturing histological features related to the underlying components of AD neuropathological severity. For CTE, predicted probabilities similarly distinguished cases without CTE from those with high-stage CTE (stages III-IV), with stronger associations for multimodal than unimodal predictions across all stain groups. The associations between predicted probabilities and neuropathological status persisted when the defining stain was withheld. For AD, all but one pairwise Reagan-category comparison remained statistically significant under masking across the three stain groups. Similarly, predicted CTE probabilities continued to distinguish cases without CTE from those with high-stage CTE under masking. Together, these findings extend the evidence of missing-stain robustness beyond categorical classification, showing that associations between model predictions and neuropathological measures can persist when a key stain is unavailable.

Spatial interpretation provided a complementary perspective on the learned representations and their behavior under missing-stain conditions. Modality-specific attention maps showed spatial patterns broadly consistent with the histological characteristics of the corresponding stains. AT8 and A*β* attention was concentrated in relatively localized regions, consistent with the focal appearance of tau- and amyloid-associated pathological features, whereas LHE attention more frequently followed the cortical gray-matter ribbon (Figure 4A). These qualitative patterns suggest that modality-specific encoders emphasize different aspects of tissue morphology, although individual attention hotspots were not independently validated against pathologist-annotated lesions. Importantly, shared-encoder class attribution showed high spatial rank correlation between full-input and defining-stain-masked conditions across the remaining stains, both in representative cases and at the population level (Figure 4B; Supplemental Table S5). This indicates that removing one stain generally pre-served the relative spatial distribution of attribution within the other stains, complementing the observed prediction-level robustness.

Our work has a few limitations. First, CTE-positive cases were available only from UNITE, whereas AD cases were represented across all three cohorts. Although cross-validation was stratified jointly by cohort and diagnostic label, the ability to generalize CTE classification to independent CTE-positive populations remains untested. Furthermore, tissue processing and neuropathological assessment protocols were standardized across the three cohorts. As a result, whether the framework generalizes to independent brain banks with different tissue sampling, staining, and processing protocols remains to be established. Second, the anatomical scope of this study was restricted to the dorsolateral frontal cortex. Neuropathological assessment of AD relies on the regional distribution of amyloid and tau pathology, while consensus criteria for CTE recommend sampling multiple brain regions given the patchy distribution of its pathognomonic lesions [27, 1]. The present findings therefore reflect only a subset of the histopathological information relevant to AD and CTE, and more comprehensive neuropathological characterization will require integration of information across multiple brain regions. Third, direct supervision on disease-specific severity measures was not investigated, and the restriction to high-stage CTE precluded assessment across the full spectrum of CTE severity. Fourth, the spatial interpretation analysis was restricted to cases with all three stains available for full-versus-masked comparisons, and attention localization was not validated against expert lesion annotations. In addition, AT8 and A*β* patches were restricted to gray matter, whereas LHE included both gray and white matter. Whether these differences in tissue coverage influence the shared encoder’s spatial attribution, particularly for LHE, remains unexamined.

In conclusion, our model’s tolerance to absent stains could facilitate retrospective re-assessment of archival brain bank material, including cases in which CTE was not evaluated during the original examination and for which additional tissue sampling or staining may no longer be feasible. Such applications can potentially expand the effective sample size available for epidemiological inference. Also, the ability to accommodate varying stain combinations provides a methodological basis for future studies on brain banks with different tissue sampling and staining protocols. Additional extensions include incorporating proteinopathy markers beyond the current panel (e.g., TDP-43, *α*-synuclein, and glial-specific stains) and external validation on independent CTE-enriched cohorts spanning a broader range of neuropathological stages.

### Disclosures

V.B.K. is the scientific founder of Cognimark, Inc. The remaining authors declare no competing interests.

### Institutional approval

All procedures complied with the Declaration of Helsinki. Institutional review board approval was obtained from Boston University School of Medicine, VA Boston Healthcare System, and VA Bedford Healthcare System. Written informed consent was obtained from the donor’s next of kin.

