## Supplement for "Stain-robust classification of Alzheimer’s disease and chronic traumatic encephalopathy"

Supplemental information

### Supplemental figure legends

**Figure S1. Segmentation model performance and comparison against Trident's built-in approach. A.** Mean Dice coefficient for the AT8 and A$\beta$ segmentation models on training and hold-out test sets. The AT8 model achieves Dice of $0.909 \pm 0.066$ (train, $n = 203$) and $0.900 \pm 0.074$ (test, $n = 49$). The A$\beta$ model achieves Dice of $0.977 \pm 0.020$ (train, $n = 175$) and $0.966 \pm 0.032$ (test, $n = 43$). **B.** Representative AT8 study-population cases where Trident's default approach (DeepLabV3 pretrained on HEST-1k) fails due to low staining intensity. Each row shows the WSI thumbnail, the tissue mask by Trident default and its thumbnail overlay, the tissue mask by the fine-tuned UNet and its thumbnail overlay (green = segmented tissue). The fine-tuned UNet recovers large tissue regions missed by the default model. **C.** Same comparison for representative A$\beta$ cases. The fine-tuned UNet consistently identifies tissue regions that the default model either omits or conflates with background.

**Figure S2. Unimodal model-predicted AD and CTE probabilities across neuropathological staging categories.** Per-subject predicted probabilities of AD and CTE are shown using pooled out-of-fold test predictions from all five cross-validation folds. **A-C.** Predicted AD probabilities across Reagan categories (no AD, low, intermediate, and high) from the unimodal models for the AT8 (**A**), A$\beta$ (**B**), and LHE (**C**) groups. **D-F.** Predicted CTE probabilities for subjects without CTE and those with CTE (stages III-IV) from the unimodal models for AT8 (**D**), A$\beta$ (**E**), and LHE (**F**). Within each panel, individual subjects are represented by scatter points (circles: UNITE; triangles: FHS; diamonds: ADRC).

Boxplots summarize the distribution within each Reagan category (center line: median; box: interquartile range; whiskers extend to the most extreme observations within 1.5$\times$ the interquartile range), and dotted grey lines connect the category medians. Pairwise differences between staging categories are assessed using two-sided Mann-Whitney $U$ tests, with Holm-Bonferroni correction across the six Reagan-category comparisons within each AD panel (A-C). CTE panels (D-F) compare the two available CTE categories. Significance is indicated by * for $p < 0.05$, ** for $p < 0.01$, *** for $p < 0.001$, and **** for $p < 0.0001$ (adjusted $p$-values for A-C). Spearman rank-order correlation coefficients ($\rho$) and their corresponding $p$-values are displayed in the upper-left corner of each panel.

| Stain group | Age (mean$_{SD}$, [min, max]) | Male (n, %) | Stains available (AT8, A$\beta$, LHE) | CTE stage (III, IV) | AD/Reagan stage (Not AD, Low, Intermediate, High) | Cohort composition (UNITE, FHS, ADRC) |
|---|---|---|---|---|---|---|
| **AT8** [n=512] | 74.16$_{14.17}$, [25, 107] | 435, 84.96% | 512, 429, 486 | 208, 124 | 220, 128, 93, 62 | 360, 78, 74 |
| Control [n=80, 15.62%] | 83.31$_{12.69}$, [51, 105] | 48, 60.00% | 80, 69, 71 | NA | 37, 42, 0, 0 | 13, 59, 8 |
| AD [n=97, 18.95%] | 82.49$_{10.87}$, [55, 107] | 52, 53.61% | 97, 69, 86 | NA | 0, 0, 48, 49 | 12, 19, 66 |
| CTE [n=277, 54.10%] | 68.50$_{13.69}$, [25, 98] | 277, 100.00% | 277, 242, 274 | 188, 87 | 183, 86, 0, 0 | 277, 0, 0 |
| CTE+AD [n=58, 11.33%] | 74.36$_{8.94}$, [54, 92] | 58, 100.00% | 58, 49, 55 | 20, 37 | 0, 0, 45, 13 | 58, 0, 0 |
| **A$\beta$** [n=519] | 74.36$_{13.62}$, [25, 106] | 461, 88.82% | 429, 519, 493 | 225, 121 | 216, 128, 102, 65 | 371, 69, 79 |
| Control [n=69, 13.29%] | 83.78$_{12.41}$, [51, 105] | 41, 59.42% | 69, 69, 60 | NA | 31, 38, 0, 0 | 11, 50, 8 |
| AD [n=102, 19.65%] | 81.61$_{9.86}$, [55, 106] | 72, 70.59% | 69, 102, 91 | NA | 0, 0, 51, 51 | 12, 19, 71 |
| CTE [n=283, 54.53%] | 69.30$_{13.61}$, [25, 98] | 283, 100.00% | 242, 283, 280 | 197, 84 | 185, 90, 0, 0 | 283, 0, 0 |
| CTE+AD [n=65, 12.52%] | 74.77$_{8.81}$, [54, 92] | 65, 100.00% | 49, 65, 62 | 28, 37 | 0, 0, 51, 14 | 65, 0, 0 |
| **LHE** [n=576] | 74.14$_{13.89}$, [25, 107] | 495, 85.94% | 486, 493, 576 | 246, 137 | 242, 134, 113, 77 | 408, 82, 86 |
| Control [n=71, 12.33%] | 83.85$_{12.38}$, [51, 105] | 39, 54.93% | 71, 60, 71 | NA | 31, 39, 0, 0 | 10, 59, 2 |
| AD [n=119, 20.66%] | 82.26$_{10.53}$, [55, 107] | 70, 58.82% | 86, 91, 119 | NA | 0, 0, 59, 60 | 12, 23, 84 |
| CTE [n=315, 54.69%] | 68.80$_{13.63}$, [25, 98] | 315, 100.00% | 274, 280, 315 | 218, 95 | 211, 95, 0, 0 | 315, 0, 0 |
| CTE+AD [n=71, 12.33%] | 74.28$_{8.75}$, [54, 91] | 71, 100.00% | 55, 62, 71 | 28, 42 | 0, 0, 54, 17 | 71, 0, 0 |

Table S1: **Study population by stain group.** Summary of demographic and clinical attributes for the three stain-defined groups (Group AT8 / A$\beta$ / LHE = all subjects with that stain available). Each group section reports the mean age (with the SD as a gray subscript and the [min, max] range), number of male subjects (with percentage), the availability of WSI stains (AT8, A$\beta$, and LHE), the CTE stage breakdown (Stage III and Stage IV), the AD/Reagan stage breakdown (Not AD, Low Probability, Intermediate Probability, High Probability), and the cohort composition (number of subjects from each of UNITE, FHS, and ADRC) of the group and each of its diagnostic subgroups. AD status is defined from the NIA-Reagan neuropathological category (Intermediate/High probability = AD-positive; Not AD/Low probability = AD-negative); one Stage II CTE case is excluded (low-grade CTE, Stage I/II, out of scope).

| Group | Condition | Fold | AUROC | AP | Accuracy | Precision | Sensitivity | Specificity |
|---|---|---|---|---|---|---|---|---|
| AT8 | Multimodal | 1 | 0.906 | 0.744 | 0.716 | 0.652 | 0.695 | 0.905 |
| | | 2 | 0.868 | 0.684 | 0.714 | 0.681 | 0.656 | 0.888 |
| | | 3 | 0.923 | 0.757 | 0.735 | 0.691 | 0.592 | 0.886 |
| | | 4 | 0.927 | 0.771 | 0.768 | 0.646 | 0.643 | 0.901 |
| | | 5 | 0.891 | 0.734 | 0.825 | 0.805 | 0.763 | 0.926 |
| AT8 | Multimodal (masked) | 1 | 0.851 | 0.659 | 0.642 | 0.571 | 0.603 | 0.881 |
| | | 2 | 0.782 | 0.547 | 0.552 | 0.448 | 0.497 | 0.836 |
| | | 3 | 0.880 | 0.638 | 0.725 | 0.639 | 0.628 | 0.898 |
| | | 4 | 0.866 | 0.670 | 0.687 | 0.581 | 0.591 | 0.883 |
| | | 5 | 0.818 | 0.597 | 0.608 | 0.435 | 0.489 | 0.854 |
| AT8 | Unimodal | 1 | 0.869 | 0.721 | 0.706 | 0.655 | 0.635 | 0.889 |
| | | 2 | 0.916 | 0.755 | 0.714 | 0.633 | 0.669 | 0.900 |
| | | 3 | 0.867 | 0.747 | 0.755 | 0.725 | 0.766 | 0.914 |
| | | 4 | 0.927 | 0.793 | 0.798 | 0.702 | 0.677 | 0.918 |
| | | 5 | 0.893 | 0.813 | 0.804 | 0.799 | 0.717 | 0.914 |
| A$\beta$ | Multimodal | 1 | 0.914 | 0.777 | 0.764 | 0.747 | 0.664 | 0.899 |
| | | 2 | 0.918 | 0.782 | 0.783 | 0.746 | 0.780 | 0.920 |
| | | 3 | 0.874 | 0.651 | 0.692 | 0.578 | 0.564 | 0.883 |
| | | 4 | 0.934 | 0.823 | 0.752 | 0.767 | 0.707 | 0.906 |
| | | 5 | 0.950 | 0.855 | 0.847 | 0.888 | 0.727 | 0.934 |
| A$\beta$ | Multimodal (masked) | 1 | 0.869 | 0.719 | 0.773 | 0.731 | 0.671 | 0.904 |
| | | 2 | 0.902 | 0.760 | 0.726 | 0.680 | 0.714 | 0.899 |
| | | 3 | 0.832 | 0.640 | 0.615 | 0.521 | 0.553 | 0.865 |
| | | 4 | 0.896 | 0.769 | 0.693 | 0.654 | 0.625 | 0.877 |
| | | 5 | 0.918 | 0.800 | 0.786 | 0.778 | 0.666 | 0.908 |
| A$\beta$ | Unimodal | 1 | 0.876 | 0.741 | 0.736 | 0.684 | 0.657 | 0.896 |
| | | 2 | 0.887 | 0.716 | 0.717 | 0.667 | 0.651 | 0.890 |
| | | 3 | 0.844 | 0.612 | 0.644 | 0.559 | 0.635 | 0.882 |
| | | 4 | 0.879 | 0.684 | 0.644 | 0.537 | 0.518 | 0.862 |
| | | 5 | 0.890 | 0.745 | 0.714 | 0.651 | 0.624 | 0.880 |

*(continued)*

| Group | Model | Fold | AUROC | AP | Accuracy | Precision | Sensitivity | Specificity |
|---|---|---|---|---|---|---|---|---|
| LHE | Multimodal | 1 | 0.902 | 0.732 | 0.700 | 0.645 | 0.679 | 0.889 |
| | | 2 | 0.910 | 0.769 | 0.746 | 0.739 | 0.744 | 0.914 |
| | | 3 | 0.889 | 0.726 | 0.783 | 0.724 | 0.691 | 0.911 |
| | | 4 | 0.918 | 0.762 | 0.761 | 0.702 | 0.738 | 0.915 |
| | | 5 | 0.912 | 0.788 | 0.764 | 0.718 | 0.684 | 0.897 |
| LHE | Multimodal (masked) | 1 | 0.899 | 0.742 | 0.658 | 0.606 | 0.659 | 0.886 |
| | | 2 | 0.919 | 0.785 | 0.763 | 0.740 | 0.776 | 0.920 |
| | | 3 | 0.878 | 0.731 | 0.730 | 0.674 | 0.680 | 0.905 |
| | | 4 | 0.924 | 0.781 | 0.752 | 0.721 | 0.727 | 0.908 |
| | | 5 | 0.886 | 0.745 | 0.709 | 0.642 | 0.678 | 0.895 |
| LHE | Unimodal | 1 | 0.852 | 0.702 | 0.742 | 0.749 | 0.619 | 0.883 |
| | | 2 | 0.858 | 0.677 | 0.669 | 0.592 | 0.620 | 0.885 |
| | | 3 | 0.872 | 0.714 | 0.748 | 0.690 | 0.654 | 0.899 |
| | | 4 | 0.847 | 0.633 | 0.690 | 0.591 | 0.577 | 0.876 |
| | | 5 | 0.827 | 0.636 | 0.691 | 0.583 | 0.588 | 0.870 |

Table S2: **Per-fold model performance on four-class classification (extends Table 3).** For each target stain (AT8 / A$\beta$ / LHE), the unimodal model (target stain only), the multimodal model (target stain plus all other available stains), and the multimodal model with the target stain masked are evaluated via stratified 5-fold cross-validation. AUROC and AP are macro-averaged across the four classes using a one-vs-rest formulation. Accuracy is calculated from arg-max predictions. Precision, sensitivity, and specificity are macro-averaged across classes. Each row reports these metrics computed independently on one fold's own held-out test set.

| Comparison | Group | $\Delta$AUROC$_{\text{SE}}$ | 95% CI | $p$-value | Adjusted $p$-value |
|---|---|---|---|---|---|
| Multimodal vs. unimodal | AT8 | $0.025_{0.011}$ | [0.003, 0.047] | $2.89 \times 10^{-2}$ | |
| | A$\beta$ | $0.045_{0.011}$ | [0.023, 0.067] | $4.56 \times 10^{-5}$ | |
| | LHE | $0.063_{0.014}$ | [0.037, 0.090] | $2.65 \times 10^{-6}$ | |
| | **Combined** | $0.044_{0.007}$ | [0.031, 0.058] | $1.61 \times 10^{-10}$ | $3.22 \times 10^{-10}$ |
| Masked multimodal vs. unimodal | AT8 | $-0.038_{0.016}$ | [−0.071, −0.006] | $1.88 \times 10^{-2}$ | |
| | A$\beta$ | $0.013_{0.015}$ | [−0.017, 0.042] | $3.97 \times 10^{-1}$ | |
| | LHE | $0.057_{0.016}$ | [0.027, 0.088] | $2.56 \times 10^{-4}$ | |
| | **Combined** | $0.011_{0.009}$ | [−0.007, 0.028] | $2.44 \times 10^{-1}$ | $2.44 \times 10^{-1}$ |
| Masked multimodal vs. multimodal | AT8 | $-0.063_{0.009}$ | [−0.081, −0.047] | $6.40 \times 10^{-13}$ | |
| | A$\beta$ | $-0.033_{0.007}$ | [−0.047, −0.019] | $8.01 \times 10^{-6}$ | |
| | LHE | $-0.006_{0.006}$ | [−0.018, 0.006] | $3.28 \times 10^{-1}$ | |
| | **Combined** | $-0.034_{0.004}$ | [−0.042, −0.026] | $2.22 \times 10^{-15}$ | $6.66 \times 10^{-15}$ |

Table S3: **Pairwise comparisons of macro-averaged four-class AUROC among unimodal, multimodal, and defining-stain-masked multimodal models.** Each row compares two models using pooled out-of-fold predictions from all five cross-validation folds within a stain group. Masked multimodal refers to the multimodal model evaluated with the group's defining stain withheld at inference. $\Delta$AUROC is calculated as the macro-averaged four-class AUROC of the first model minus that of the second (Table 3), with bootstrap standard errors displayed as grey subscripts. Standard errors and 95% confidence intervals are estimated from 10,000 class-stratified bootstrap resamples of subjects, with paired model predictions preserved within each resample. Two-sided $p$-values are calculated using Wald tests based on the ratio of $\Delta$AUROC to its bootstrap standard error and the standard normal approximation. Combined estimates are obtained by averaging the three stain groups' $\Delta$AUROC values and corresponding bootstrap distributions with equal weight per group. Combined $p$-values are Holm-Bonferroni-adjusted across the three pairwise model comparisons. Group-level $p$-values are unadjusted.

| Model | Group | Comparison | $U$ | $p$-value | Adjusted $p$-value |
|---|---|---|---|---|---|
| Unimodal | AT8 | no AD vs low | 11268 | $1.89 \times 10^{-3}$ | $1.89 \times 10^{-3}$ |
| | | no AD vs intermediate | 2546 | $8.54 \times 10^{-26}$ | $4.27 \times 10^{-25}$ |
| | | no AD vs high | 460 | $3.53 \times 10^{-29}$ | $2.12 \times 10^{-28}$ |
| | | low vs intermediate | 2428 | $5.99 \times 10^{-14}$ | $1.80 \times 10^{-13}$ |
| | | low vs high | 643 | $8.44 \times 10^{-21}$ | $3.37 \times 10^{-20}$ |
| | | intermediate vs high | 1768 | $4.69 \times 10^{-5}$ | $9.37 \times 10^{-5}$ |
| Unimodal | A$\beta$ | no AD vs low | 9029 | $7.55 \times 10^{-8}$ | $1.51 \times 10^{-7}$ |
| | | no AD vs intermediate | 1591 | $7.54 \times 10^{-35}$ | $4.53 \times 10^{-34}$ |
| | | no AD vs high | 504 | $8.03 \times 10^{-30}$ | $4.01 \times 10^{-29}$ |
| | | low vs intermediate | 3095 | $7.55 \times 10^{-12}$ | $2.26 \times 10^{-11}$ |
| | | low vs high | 1190 | $5.64 \times 10^{-16}$ | $2.26 \times 10^{-15}$ |
| | | intermediate vs high | 2308 | $9.54 \times 10^{-4}$ | $9.54 \times 10^{-4}$ |
| Unimodal | LHE | no AD vs low | 13628 | $1.04 \times 10^{-2}$ | $1.04 \times 10^{-2}$ |
| | | no AD vs intermediate | 4327 | $3.20 \times 10^{-25}$ | $1.60 \times 10^{-24}$ |
| | | no AD vs high | 1187 | $9.04 \times 10^{-31}$ | $5.42 \times 10^{-30}$ |
| | | low vs intermediate | 3106 | $1.45 \times 10^{-15}$ | $4.36 \times 10^{-15}$ |
| | | low vs high | 921 | $3.24 \times 10^{-23}$ | $1.30 \times 10^{-22}$ |
| | | intermediate vs high | 2432 | $2.55 \times 10^{-7}$ | $5.10 \times 10^{-7}$ |
| Multimodal | AT8 | no AD vs low | 9337 | $1.60 \times 10^{-7}$ | $3.20 \times 10^{-7}$ |
| | | no AD vs intermediate | 1290 | $2.50 \times 10^{-34}$ | $1.50 \times 10^{-33}$ |
| | | no AD vs high | 235 | $3.69 \times 10^{-31}$ | $1.84 \times 10^{-30}$ |
| | | low vs intermediate | 2176 | $8.60 \times 10^{-16}$ | $2.58 \times 10^{-15}$ |
| | | low vs high | 661 | $1.36 \times 10^{-20}$ | $5.44 \times 10^{-20}$ |
| | | intermediate vs high | 1848 | $1.58 \times 10^{-4}$ | $1.58 \times 10^{-4}$ |
| Multimodal | A$\beta$ | no AD vs low | 9143 | $1.52 \times 10^{-7}$ | $3.05 \times 10^{-7}$ |
| | | no AD vs intermediate | 1155 | $5.50 \times 10^{-38}$ | $3.30 \times 10^{-37}$ |
| | | no AD vs high | 98 | $1.94 \times 10^{-33}$ | $9.71 \times 10^{-33}$ |
| | | low vs intermediate | 2131 | $1.79 \times 10^{-18}$ | $5.37 \times 10^{-18}$ |
| | | low vs high | 521 | $3.38 \times 10^{-23}$ | $1.35 \times 10^{-22}$ |
| | | intermediate vs high | 2150 | $1.32 \times 10^{-4}$ | $1.32 \times 10^{-4}$ |
| Multimodal | LHE | no AD vs low | 10422 | $9.59 \times 10^{-9}$ | $1.92 \times 10^{-8}$ |
| | | no AD vs intermediate | 1906 | $5.31 \times 10^{-39}$ | $3.19 \times 10^{-38}$ |
| | | no AD vs high | 420 | $1.62 \times 10^{-36}$ | $8.12 \times 10^{-36}$ |
| | | low vs intermediate | 2827 | $2.26 \times 10^{-17}$ | $6.78 \times 10^{-17}$ |

*(continued)*

| Model | Group | Comparison | $U$ | $p$-value | Adjusted $p$-value |
|---|---|---|---|---|---|
| | | low vs high | 881 | $1.26 \times 10^{-23}$ | $5.04 \times 10^{-23}$ |
| | | intermediate vs high | 2702 | $9.49 \times 10^{-6}$ | $9.49 \times 10^{-6}$ |
| Multimodal (masked) | AT8 | no AD vs low | 9345 | $1.68 \times 10^{-7}$ | $3.36 \times 10^{-7}$ |
| | | no AD vs intermediate | 2323 | $3.23 \times 10^{-27}$ | $1.94 \times 10^{-26}$ |
| | | no AD vs high | 820 | $3.77 \times 10^{-26}$ | $1.88 \times 10^{-25}$ |
| | | low vs intermediate | 2779 | $1.38 \times 10^{-11}$ | $4.13 \times 10^{-11}$ |
| | | low vs high | 1131 | $1.45 \times 10^{-15}$ | $5.81 \times 10^{-15}$ |
| | | intermediate vs high | 2176 | $9.87 \times 10^{-3}$ | $9.87 \times 10^{-3}$ |
| Multimodal (masked) | A$\beta$ | no AD vs low | 12103 | $5.36 \times 10^{-2}$ | $5.36 \times 10^{-2}$ |
| | | no AD vs intermediate | 2782 | $5.40 \times 10^{-27}$ | $2.70 \times 10^{-26}$ |
| | | no AD vs high | 583 | $3.83 \times 10^{-29}$ | $2.30 \times 10^{-28}$ |
| | | low vs intermediate | 2524 | $1.40 \times 10^{-15}$ | $4.19 \times 10^{-15}$ |
| | | low vs high | 704 | $4.43 \times 10^{-21}$ | $1.77 \times 10^{-20}$ |
| | | intermediate vs high | 1957 | $8.36 \times 10^{-6}$ | $1.67 \times 10^{-5}$ |
| Multimodal (masked) | LHE | no AD vs low | 10730 | $5.55 \times 10^{-8}$ | $1.11 \times 10^{-7}$ |
| | | no AD vs intermediate | 2175 | $2.57 \times 10^{-37}$ | $1.54 \times 10^{-36}$ |
| | | no AD vs high | 511 | $8.30 \times 10^{-36}$ | $4.15 \times 10^{-35}$ |
| | | low vs intermediate | 2616 | $8.24 \times 10^{-19}$ | $2.47 \times 10^{-18}$ |
| | | low vs high | 697 | $1.47 \times 10^{-25}$ | $5.88 \times 10^{-25}$ |
| | | intermediate vs high | 2759 | $1.91 \times 10^{-5}$ | $1.91 \times 10^{-5}$ |

Table S4: **Pairwise comparisons of predicted AD probabilities across Reagan categories (extends Figure 3 and Supplemental Figure S2).** For each stain group and model condition, all six pairwise comparisons between Reagan categories are assessed using two-sided Mann-Whitney $U$ tests on pooled out-of-fold predictions from all five cross-validation folds. Holm-Bonferroni correction is applied across the six comparisons within each model–group combination. Both unadjusted and adjusted $p$-values are reported.

| Group | Non-defining stain | Ground-truth label | $N$ | $\text{Mean}_{\text{SD}}$ | Median | $p$-value | Adjusted $p$-value |
|---|---|---|---|---|---|---|---|
| AT8 | A$\beta$ | Control | 60 | $0.830_{0.150}$ | 0.878 | $1.85 \times 10^{-2}$ | $1.85 \times 10^{-2}$ |
| | | AD | 58 | $0.952_{0.051}$ | 0.970 | $6.78 \times 10^{-12}$ | $1.22 \times 10^{-10}$ |
| | | CTE | 239 | $0.834_{0.145}$ | 0.877 | $2.93 \times 10^{-10}$ | $2.93 \times 10^{-9}$ |
| | | CTE+AD | 46 | $0.940_{0.052}$ | 0.950 | $1.95 \times 10^{-11}$ | $3.31 \times 10^{-10}$ |
| | LHE | Control | 60 | $0.957_{0.035}$ | 0.964 | $2.71 \times 10^{-11}$ | $3.84 \times 10^{-10}$ |
| | | AD | 58 | $0.963_{0.046}$ | 0.979 | $6.14 \times 10^{-12}$ | $1.17 \times 10^{-10}$ |
| | | CTE | 239 | $0.832_{0.136}$ | 0.867 | $3.65 \times 10^{-9}$ | $2.56 \times 10^{-8}$ |
| | | CTE+AD | 46 | $0.931_{0.072}$ | 0.963 | $2.03 \times 10^{-10}$ | $2.23 \times 10^{-9}$ |
| A$\beta$ | AT8 | Control | 60 | $0.903_{0.110}$ | 0.931 | $8.76 \times 10^{-8}$ | $4.38 \times 10^{-7}$ |
| | | AD | 58 | $0.933_{0.072}$ | 0.955 | $4.96 \times 10^{-11}$ | $5.95 \times 10^{-10}$ |
| | | CTE | 239 | $0.876_{0.104}$ | 0.907 | $4.16 \times 10^{-22}$ | $9.57 \times 10^{-21}$ |
| | | CTE+AD | 46 | $0.914_{0.081}$ | 0.935 | $1.01 \times 10^{-9}$ | $8.07 \times 10^{-9}$ |
| | LHE | Control | 60 | $0.894_{0.138}$ | 0.958 | $4.39 \times 10^{-6}$ | $8.78 \times 10^{-6}$ |
| | | AD | 58 | $0.972_{0.032}$ | 0.977 | $5.84 \times 10^{-12}$ | $1.17 \times 10^{-10}$ |
| | | CTE | 239 | $0.834_{0.151}$ | 0.881 | $2.76 \times 10^{-7}$ | $1.10 \times 10^{-6}$ |
| | | CTE+AD | 46 | $0.956_{0.048}$ | 0.972 | $2.40 \times 10^{-11}$ | $3.84 \times 10^{-10}$ |
| LHE | A$\beta$ | Control | 60 | $0.880_{0.096}$ | 0.904 | $9.53 \times 10^{-7}$ | $2.86 \times 10^{-6}$ |
| | | AD | 58 | $0.976_{0.028}$ | 0.984 | $5.56 \times 10^{-12}$ | $1.17 \times 10^{-10}$ |
| | | CTE | 239 | $0.851_{0.116}$ | 0.888 | $2.08 \times 10^{-13}$ | $4.58 \times 10^{-12}$ |
| | | CTE+AD | 46 | $0.966_{0.050}$ | 0.978 | $2.53 \times 10^{-11}$ | $3.84 \times 10^{-10}$ |
| | AT8 | Control | 60 | $0.929_{0.080}$ | 0.955 | $5.65 \times 10^{-10}$ | $5.08 \times 10^{-9}$ |
| | | AD | 58 | $0.923_{0.118}$ | 0.970 | $6.36 \times 10^{-8}$ | $3.81 \times 10^{-7}$ |
| | | CTE | 239 | $0.885_{0.104}$ | 0.925 | $3.36 \times 10^{-22}$ | $8.06 \times 10^{-21}$ |
| | | CTE+AD | 46 | $0.955_{0.061}$ | 0.980 | $3.27 \times 10^{-11}$ | $4.26 \times 10^{-10}$ |
| **Overall** | | | 2418 | $0.887_{0.120}$ | 0.927 | $4.96 \times 10^{-212}$ | $1.24 \times 10^{-210}$ |

Table S5: **Population-level spatial attribution consistency between full and defining-stain-masked multimodal models (extends Figure 4B).** For subjects with all three stains available, Spearman correlations ($\rho$) between full and defining-stain-masked shared-encoder attribution vectors are calculated for each non-defining stain using the true diagnostic label as the attribution target. Analyses use pooled out-of-fold evaluations across five cross-validation folds. Sample sizes ($N$), means with standard deviations (SD), and medians are reported by stain group, non-defining stain, and diagnostic label. The Overall row pools 2,418 subject–stain–label observations. One-sided Wilcoxon signed-rank tests assess whether median $\rho > 0.8$, with Holm-Bonferroni correction across 25 tests. Unadjusted and adjusted $p$-values are reported.

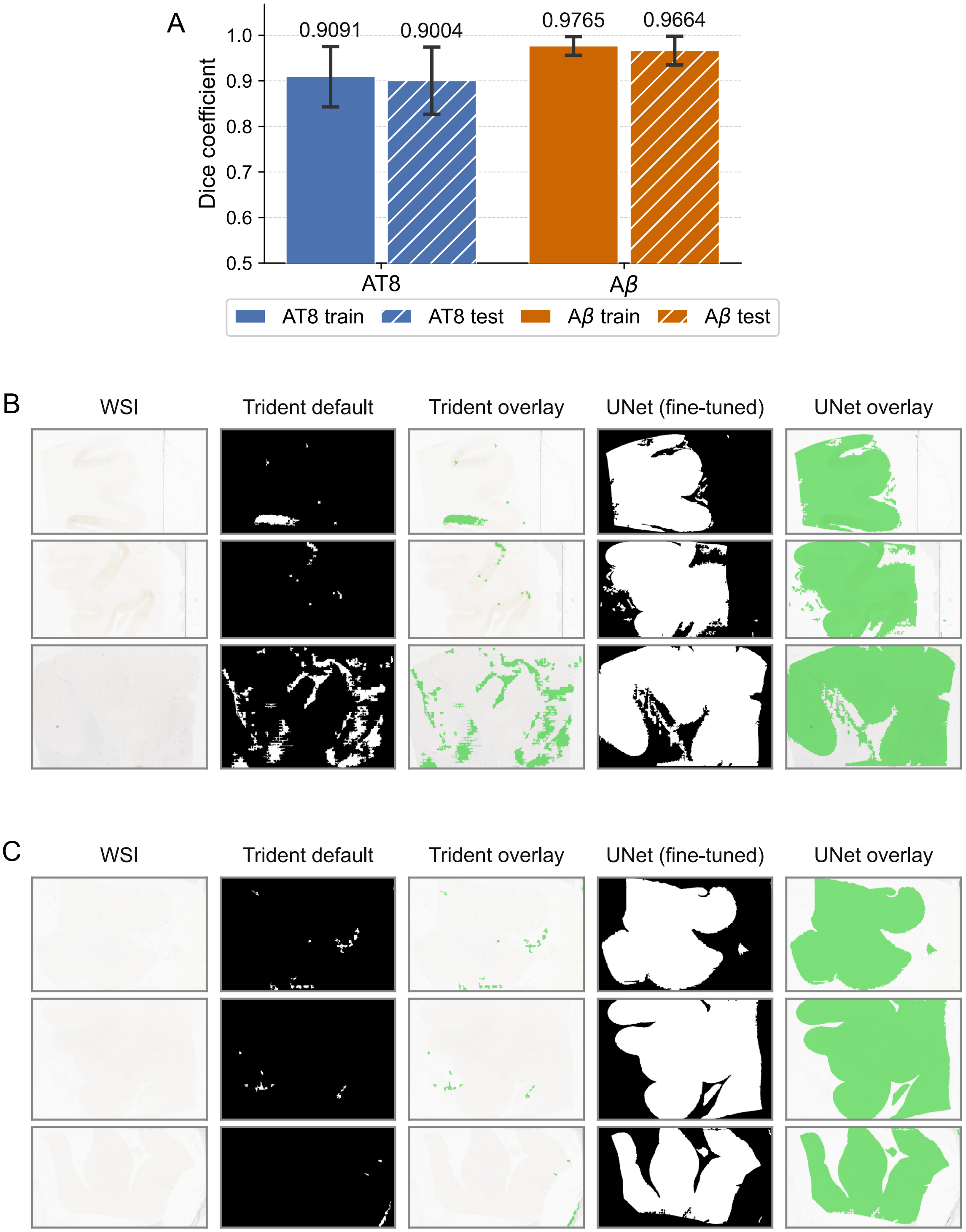
A
Dice coefficient
0.9091
0.9004
0.9765
0.9664
AT8
Aβ
AT8 train
AT8 test
Aβ train
Aβ test
B
WSI
Trident default
Trident overlay
UNet (fine-tuned)
UNet overlay
C
WSI
Trident default
Trident overlay
UNet (fine-tuned)
UNet overlay

Figure S1

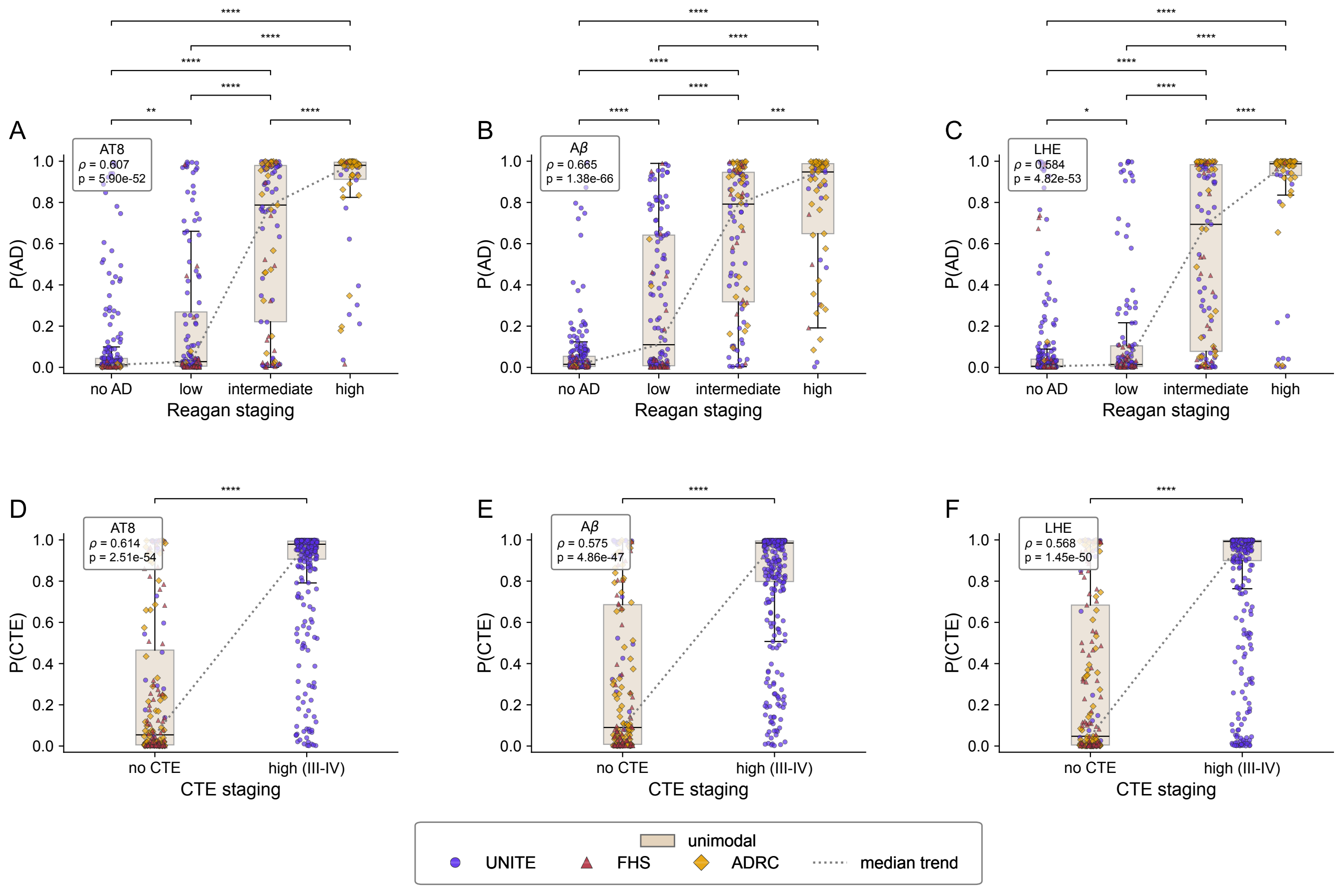

A
AT8
ρ = 0.607
p = 5.90e-52
B
Aβ
ρ = 0.665
p = 1.38e-66
C
LHE
ρ = 0.584
p = 4.82e-53
P(AD)
no AD
low
intermediate
high
Reagan staging
D
AT8
ρ = 0.614
p = 2.51e-54
E
Aβ
ρ = 0.575
p = 4.86e-47
F
LHE
ρ = 0.568
p = 1.45e-50
P(CTE)
no CTE
high (III-IV)
CTE staging
unimodal
UNITE
FHS
ADRC
median trend


Figure S2
